# mtDNA copy number, mtDNA total somatic deletions, Complex I activity, synapse number and synaptic mitochondria number are altered in schizophrenia and bipolar disorder

**DOI:** 10.1101/2022.03.04.22271871

**Authors:** Sujan C. Das, B.E. Hjelm, B. Galke, L. Morgan, A.A. Omidsalar, A. Sequeira, A.F. Schatzberg, J.D. Barchas, F.S. Lee, R.M. Myers, S.J. Watson, H. Akil, W.E. Bunney, M.P. Vawter

**Author notes:** Corresponding Author: Address: Marquis P. Vawter, Department of Psychiatry & Human Behavior. 837 Health Sciences Rd., Gillespie Neuroscience, Room 2119 University of California, Irvine - College of Medicine Irvine, CA 92697-4260.

## Abstract

Mitochondrial dysfunction is a neurobiological phenomenon implicated in the pathophysiology of schizophrenia and bipolar disorder that can synergistically affect synaptic neurotransmission. We hypothesized that schizophrenia and bipolar disorder share molecular alterations at the mitochondrial and synaptic level: mtDNA copy number (CN), mtDNA common deletion (CD), mtDNA total deletion, complex I activity, synapse number, and synaptic mitochondria number. These mitochondrial parameters were studied in dorsolateral prefrontal cortex (DLPFC), superior temporal gyrus (STG), primary visual cortex (V1), and nucleus accumbens (NAc) of 44 controls (CON), 27 schizophrenia (SZ), and 26 bipolar disorder (BD) subjects. The results showed- (i) the mtDNA CN is significantly higher in DLPFC of both SZ and BD, decreased in the STG of BD, and unaltered in V1 and NAc of both SZ and BD; (ii) the mtDNA CD is significantly higher in DLPFC of BD while unaltered in STG, V1 and NAc of both SZ and BD; (iii) The total deletion burden is significantly higher in DLPFC in both SZ and BD while unaltered in STG, V1, and NAc of SZ and BD; (iv) Complex I activity is significantly lower in DLPFC of both SZ and BD with no alteration in STG, V1 and NAc. (v) The number of synapses is decreased in DLPFC of both SZ and BD, while the synaptic mitochondria number was significantly lower in female SZ and female BD compared to female controls. Overall, these findings will pave the way to better understand the pathophysiology of schizophrenia and bipolar disorder for therapeutic interventions.

## Introduction

Mitochondria are the primary energy-producers of the cells as they generate ∼90% of cell energy in the form of adenosine triphosphate (ATP). Energy production of mitochondria in the brain is critical, as the brain uses over 20% of the energy produced in the body, yet it is only about 2% of adult body weight. Mitochondria generate ATP through a complex set of enzymes called electron transport chain (ETC). The ETC consists of five protein complexes (Complex I, II, III, IV and V). The ETC proteins are encoded by two distinct genomes: the mitochondrial genome (mtDNA) and the nuclear genome (nDNA). mtDNA are closed circle, intron-free, and polyploid double-stranded DNA molecules consisting of 16569 nucleotides that encode for 13 proteins, 22 transfer RNAs and 2 ribosomal RNAs. Conversely, nDNA encodes for approximately 1500 genes involved in mitochondrial functions and localization [1]. Within ETC, Complex I is the largest multimeric protein complex and consists of 45 subunits, from which 6 subunits are encoded by mtDNA.

Schizophrenia (SZ) and bipolar disorder (BD) are complex polygenic neuropsychiatric disorders. One pathway implicated in the pathophysiology of SZ and BD is mitochondria dysfunction [1–3]. Clinical studies have shown that patients with SZ and BD have abnormal energy metabolites and redox states implicating dysfunctional mitochondria [4–6]. Genome-wide association studies (GWASs) and gene-set based analysis have linked the pathophysiology of SZ and BD to synaptic dysfunction and nuclear-encoded mitochondria genes (NEMG) [7]. Mitochondrial dysfunction in SZ and BD appears to stem from a complex set of causalities which include but are not limited to -a) single nucleotide polymorphism (SNP) in mtDNA and NEMGs [8]; b) decreased expression of genes responsible for mitochondrial function revealed by transcriptomics analysis [9, 10]; c) altered mtDNA deletion burdens in SZ and BD brain tissues [11, 12]; d) altered mtDNA copy number [13]; e) altered enzymatic activity of ETC proteins [14]; f) an altered number of mitochondria puncta and mitochondria volume density [1]. Based on above mentioned etiology, it is well established that genetic risk component plays a major role in dysfunctional mitochondria of SZ and BD. Induced pluripotent stem cells (iPSCs) derived neurons and cerebral organoids from patients with SZ and BD showed dysfunctional mitochondria which further strengthen the role of genetic risk component in pathophysiology of SZ and BD [15, 16].

Due to the complex nature of SZ and BD, we conducted a systematic investigation of critical mitochondria parameters related to mtDNA, as well as mitochondrial distribution and function, in SZ and BD brains. Specifically, we investigated the following mitochondrial parameters in clinically well-defined SZ and BD subjects: mtDNA copy number, mtDNA common deletions, mtDNA total deletions, Complex I activity, total mitochondria number, total synapse number and synaptic mitochondria number. Models tested for diagnostic effect included potential factors such as age, sex and antipsychotic medication(s). To achieve this goal, we used postmortem brain tissues from 44 control (CON), 27 schizophrenia (SZ) and 26 bipolar disorder (BD) subjects. Four key brain regions across this cohort have been investigated to identify the regional effect and specificity of any observed mitochondrial difference: dorsolateral prefrontal cortex (DLPFC), superior temporal gyrus (STG), primary visual cortex (V1) and nucleus accumbens (NAc). **First**, we determined how mtDNA copy number vary in SZ and BD compared to CON. **Second**, we measured mtDNA common deletion in SZ and BD compared to CON. **Third**, we determined the pooled effect of total deletions through our high-throughput and high-resolution pipeline called “Splice-Break” in CON, SZ and BD. **Fourth,** we determined the alteration of Complex I activity in SZ and BD. **Finally**, we calculated the overall mitochondria number, synapse number and synaptic mitochondria number through confocal microscopy and IMARIS modeling in CON, SZ and BD.

## Materials and Methods

### Subjects

Postmortem brain collection and preservation were conducted as previously published procedures [17]. Briefly, postmortem brain tissues were obtained from the UCI-Pritzker brain bank. Written consent was obtained from the next-of-kin for each subject. The postmortem brain collection and experimental procedures were approved by UCI Institutional Review Board (IRB). Psychological autopsies were conducted based on family informant interviews, subjects’ medication history, medical and psychiatric records, and coroners’ toxicology reports. Based on psychological autopsies, three clinically well-defined experimental groups were studied: Control (CON), Schizophrenia (SZ) and Bipolar Disorder (BD). In total, 44 CON, 27 SZ and 26 BD subjects were included in this study. Results for some of these parameters were previously published for DLPFC (parameters 1A, 2A, and 4A) with the same subjects [13], and are included for comparisons to three additional brain regions. The full demographics of all the subjects has been depicted in Table ST1.

### mtDNA Copy Number and Common Deletion

mtDNA copy number (mtDNA CN), mtDNA common deletion (mtDNA CD; adjusted breakpoints:8482-13460) and mtDNA amplicon resequencing was performed on extracted genomic DNA (gDNA) of samples as described in our previously published procedures [12, 13]. The in-detail procedure has been described in Supplementary Information.

### Pooled Effect of Total Deletion of mtDNA by High-resolution Pipeline “Splice-Break”

With our recently developed pipeline [18], called Splice-Break, we measured total deletions of mitochondria DNA (common deletion + other deletions) which occur at different rates in different brain regions and increases with age. The full description of the “Splice-Break” method has been explained in Supplementary Information. For more details about the “Splice-Break”, please read [18].

### Complex I Activity (activity/unit of Complex I protein)

The Complex I activity was measured based on our previously published two-steps procedure [13]. For more of this procedure, please read Supplementary Information.

### Immunohistochemistry (IHC) for Colocalization of Mitochondria and Synaptic Marker

The gene TOMM40 (translocase of outer mitochondrial membrane 40) codes for the protein TOM40 which is required for the import of proteins into the mitochondria. The PSD95 protein

(postsynaptic density protein 95) is exclusively localized in excitatory synapses. The colocalization of TOM40 and PSD95 was evaluated by immunohistochemistry (IHC) as described previously with slight modification [19]. Please read Supplementary Information for more details.

### Statistical Analyses

The raw data for each dependent variable was imported into JMP Pro 16.0 (SAS Institute, Inc., NC, USA) and checked for outliers greater than 3 standard deviations from the mean, which were removed. Additional variables beyond diagnosis and brain region (age, PMI, pH, sex) were checked for correlations with raw data and were included in models when significant. Linear regression model for single variables were used to determine group differences in a single brain region. When similar technology was used across three cortical regions in which all subjects were assayed, a repeated measure (mixed effects linear model) based on subject, region, and the diagnosis was calculated to increase the analysis robustness and compared with single region analysis. Post hoc analysis of group effects were conducted after main effects or interaction effects were determined significant (p < 0.05). Exploratory analysis of medication effects using previous grouping of subjects with neuroactive compounds in cerebellar toxicology reports from NMS Labs (PA, USA) was used as a grouping variable in pooled BD and SZ subjects, details of these subjects’ toxicology were previously published [13].

## Results

### mtDNA Copy Number in SZ and BD

Different tissues have various bioenergetics profiles which makes mtDNA copy number (CN) vary in a tissue-specific (and sometimes age-dependent) manner. Decreased peripheral mtDNA CN has been reported in SZ and BD [20, 21]. Peripheral mtDNA CN has also been shown to decrease with aging, although brain tissue mtDNA CN showed no age-related changes [22, 23]. Here, we sought to determine how SZ and BD affect the mtDNA CN in four critical brain regions of neocortex. A mixed model effects analysis, with adjustments of covariates (∼age+sex+region+subject), was conducted for mtDNA CN in the DLPFC and STG regions as a repeated analysis. The V1 and NAc regions were analyzed separately because V1 data were obtained through ddPCR and NAc regions had subset of samples. For the DLPFC, both SZ and BD groups displayed significant increases (p= 0.003 and p = 0.011, respectively) in the level of mtDNA copy number compared to CON (Figure 1A). The same subjects’ STG region showed a significant decrease (p = 0.036) in mtDNA CN in BD compared to controls and a non-significant decrease in mtDNA CN in SZ compared to controls (Figure 1B-D) (Table ST2). Although the interaction of brain region (DLPFC, STG) and diagnosis was significant (p = 0.0015), but the mtDNA copy number brain region correlations (DLPFC vs STG) were not significant (Figure SF1). In separate analyses of V1 cortex and nucleus accumbens regions, there were no significant differences between groups for mtDNA CN in the V1 (p = 0.72) or the NAc (p = 0.30). Next, we determined the effect of age on mtDNA CN in DLPFC, STG and V1. Age did not have any significant effect on mtDNA CN in DLPFC, STG and V1 (Table ST7). Another potential confounding factor affecting mtDNA CN in SZ and BD is the use of antipsychotic medications. Antipsychotic medications have been shown to decrease peripheral mtDNA CN [24]. Thus, we also tested whether antipsychotic medications had any effect on mtDNA CN or not in four brain regions included in this study. Interestingly, we did not find any medication effect on mtDNA CN in DLPFC, STG, V1 and NAc of SZ and BD patients (Table ST8), suggesting these brain regions may have more stable mitochondrial CN than blood when exposed to environmental factors or medication.

**Figure 1:**
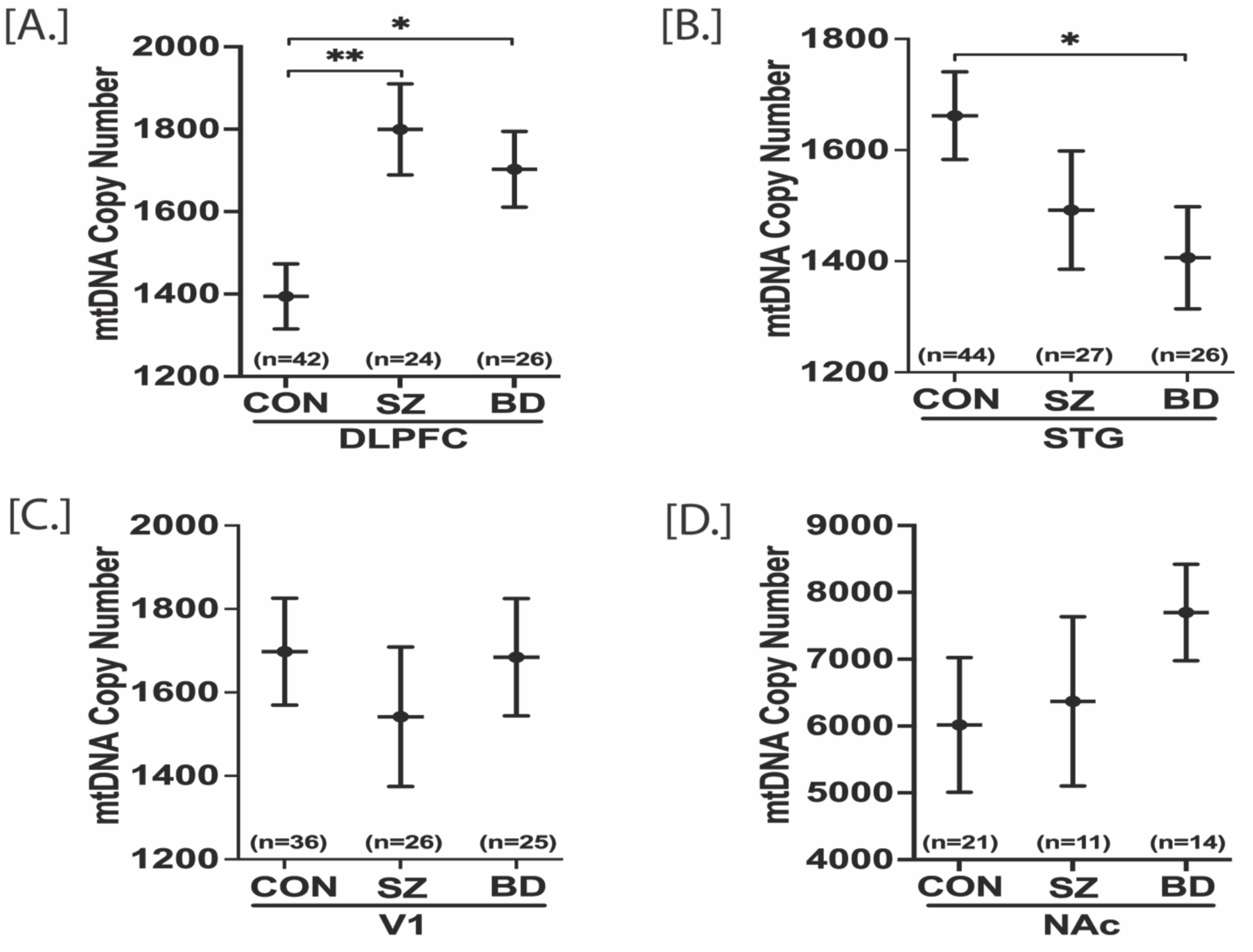
mtDNA copy number in postmortem DLPFC, STG, V1 and NAc of control (CON), schizophrenia (SZ) and bipolar disorders (BD) groups. mtDNA copy number (CN) for DLPFC, STG and NAc were obtained through qPCR while V1 mtDNA copy number (CN) was obtained through ddPCR. mtDNA CN of a sample was calculated as mtDNA CN/ALB CN. Mixed model analysis was performed for statistical significance with adjustment of age and sex. DLPFC and STG were analyzed together to obtain the regional correction whereas V1 and NAc data were analyzed individually. DLPFC mtDNA CN was significantly higher in both SZ and BD compared to control group [A]. In STG, BD group had significantly lower mtDNA CN compared to control [B]. In V1 and NAc, there were no significant groups differences [C, D]. All data are represented as least square mean (LSM) ± Std. error. p<0.05 and p<0.01 are denoted by * and **, respectively, otherwise p>0.05.

### Common Deletion of mtDNA in SZ and BD

The mtDNA common deletion (CD) is a large (4977 bp) deletion of mtDNA, shown by us and others to increase with age in multiple brain regions [12, 25]. The mtDNA CD is an indicator of oxidative stress due to ROS and the loss of 12 essential mitochondrial genes within the deleted region. mtDNA is near the electron transport chain, a major source of reactive oxygen species that can damage mtDNA and trigger deletions. Mixed model analysis was performed for statistical significance with adjustment of age and sex with the DLPFC, STG and V1 analyzed together whereas NAc data were analyzed individually. The regional mtDNA CD was increased in BD compared to CON (p =0.014) and increased in BD compared to SZ (p =0.046) in the DLPFC (Figure 2A). The effect was mainly due to increased CD in DLPFC for BD. The %CD in STG, V1 and NAc was not significantly altered (Figure 2B-D) (Table ST3). In contrast to the mtDNA CN results, the mtDNA CD showed highly significant correlations in the levels between brain regions. Regional analyses of the CD data assayed by qPCR showed a significant age effect (%CD increases with age) in each of the three cortical regions (Figure SF3). The positive correlation of age and the CD was significant in V1 cortex (p =0.032, r = 0.22), DLPFC (p = 0.0008, r = 0.34), and STG (p =0.008, r = 0.27) (Table ST9). The positive correlations of the mtDNA CD were highly significant across regions by subject, e.g., a high inter-regional correlation (STG-DLPFC, r = 0.75, 3.25E-18), which suggests a possible global mechanism for the common deletion dependent upon age (Figure SF2). A subset of subjects was assayed for the mtDNA CD in the nucleus accumbens and the inter-regional correlations remained significant when analyzing only this subset of subjects across 4 brain regions with common subjects assayed in all four regions. Further, we addressed if correlations of the mtDNA CD were altered by antipsychotic drugs and medications, for this, patients were grouped by detectable or not detectable toxicological psychotropic drugs and medications based on postmortem toxicology analysis. The inter-regional correlations remained significant for both groups regardless of toxicology measures. Thus, presence of medications or drugs did not alter the significant inter regional correlations of mtDNA CD levels, suggesting it either had no effect, or had an equivalent and non-significant effect on all brain regions we evaluated (data not shown).

**Figure 2:**
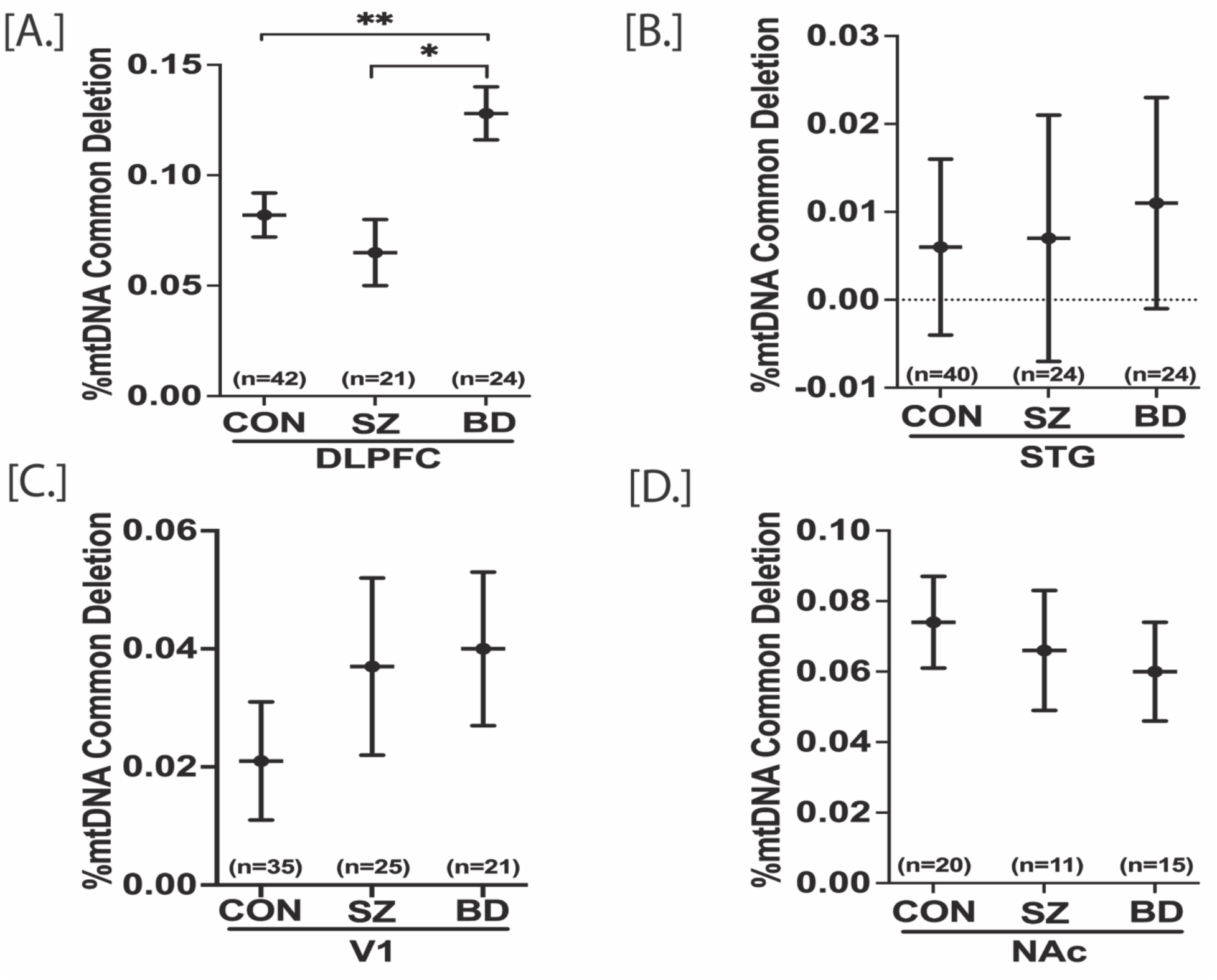
mtDNA common deletions in postmortem DLPFC, STG, V1 and NAc of control (CON), schizophrenia (SZ) and bipolar disorders (BD) groups. mtDNA common deletion (CD) for DLPFC, STG and NAc were obtained through qPCR while V1 mtDNA copy number was obtained through ddPCR. %mtDNA CD was calculated as [mtDNA CD copies/ (wild type mtDNA copies+ mtDNA CD copies)] x 100. Mixed model analysis was performed for statistical significance with adjustment of age and sex. DLPFC, STG and V1 were analyzed together to obtain the regional correction whereas NAc data were analyzed individually. DLPFC mtDNA CD was significantly higher in BD compared to control group [A]. There were no significant groups differences in STG, V1 and NAc mtDNA CD [B, C, D]. All data are represented as least square mean (LSM) ± Std. error. p<o.o5 and p<0.01 are denoted by * and **, respectively, otherwise p>0.05.

### The pooled effect of total mtDNA deletions in SZ and BD

We utilized the highly sensitive “Splice-Break” pipeline to detect the total number of unique mtDNA deletions whose breakpoints fell within the position of mtDNA 357-15925. The “Splice-Break” pipeline provided us deletion read %’s for hundreds to thousands of mtDNA deletion breakpoints, and we calculated the cumulative deletion metrics as previously described [18]. Here, we focused on the cumulative analysis of “deletions per 10K coverage”, which is the estimation of the pooled effect of total deletions species (i.e., unique set of breakpoints), as the other two metrics described in our method paper were not significantly different in group comparisons. Previously, we showed that deletions per 10K coverage parameter did not significantly vary with age, brain pH and postmortem interval (PMI) [18]. Here, we computed standardized residuals for deletions per 10K coverage with benchmark coverage and age added as a covariate. The repeated measure analysis revealed that the standardized residuals of deletions per 10K was significantly higher in DLPFC of both SZ (p=0.036) and BD (p=0.000055) compared to CON (Figure 3A). There were no other significant group comparisons within STG [CON vs SZ, p=0.78; CON vs BD, p=0.68], V1 [CON vs SZ, p=0.87; CON vs BD, p=0.71] and NAc [CON vs SZ, p=0.77; CON vs BD, p=0.40] (Figure 3B-D) (Table ST4). The deletions per 10K coverage was not significantly correlated with age (Table ST10). The medications did not affect the total deletions per 10K coverage in DLPFC, STG and V1 (Figure SF4).

**Figure 3:**
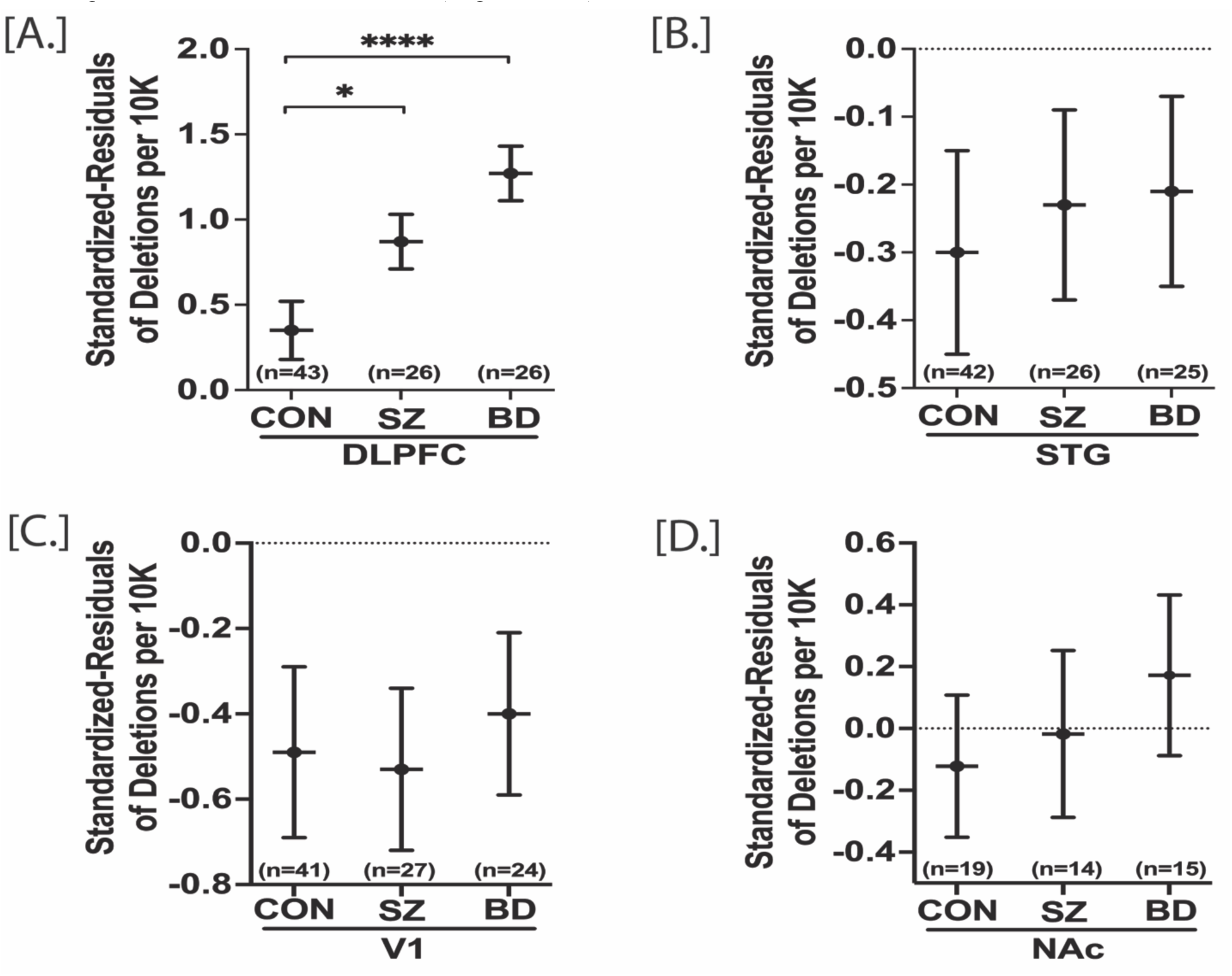
Standardized residuals of deletions per 10K in postmortem DLPFC, STG, V1 and NAc of control (CON), schizophrenia (SZ) and bipolar disorders (BD) groups. Mixed model analysis was performed for statistical significance with adjustment of age and sex. DLPFC, STG and V1 were analyzed together to obtain the regional correction whereas NAc data were analyzed individually. DLPFC standardized residuals of deletion per 10K was significantly higher in both SZ and BD compared to control group [A]. There were no significant groups differences in STG, V1 and NAc [B, C, D]. All data are represented as least square mean (LSM) ± Std. error. p<0.05 and p<0.0001 are denoted by * and ****, respectively, otherwise p>0.05.

### Complex I activity in SZ and BD

Complex I (NADH ubiquinone oxidoreductase) is the first enzyme complex for cellular oxidative phosphorylation and a major contributor to the proton gradient across the electron transport chain. It has been shown that mitochondria quality decreases with aging which in turn decreases the Complex I activity in various tissues including frontal cortex and muscle [13, 26]. Here, we sought to determine how schizophrenia and bipolar disorder affect the Complex I activity in four brain regions with adjustment of age and sex. Repeated measure analysis was performed with DLPFC, STG and V1 regions together while NAc data was analyzed separately. The effect of medication was analyzed for each four brain regions individually. The Complex I activity is significantly decreased in DLPFC of both SZ (p=0.028) and BD (p=0.012) groups compared to CON. SZ and BD did not affect the Complex I activity in STG, V1 and NAc (Figure 4) (Table ST5). The regional correlation of Complex I activity was not significant across DLPFC, STG, V1 and NAc (Table ST11). The STG Complex I activity was dependent on age while DLPFC, V1 and NAc Complex I activity was independent of age (Table ST11). An analysis of medications/drug effects on Complex I showed the DLPFC Complex I activity was affected by medications while there was no medications effect on STG, V1 and NAc Complex I activity (Table ST12). Thus, the effects of medication/drugs may be regionally specific.

**Figure 4:**
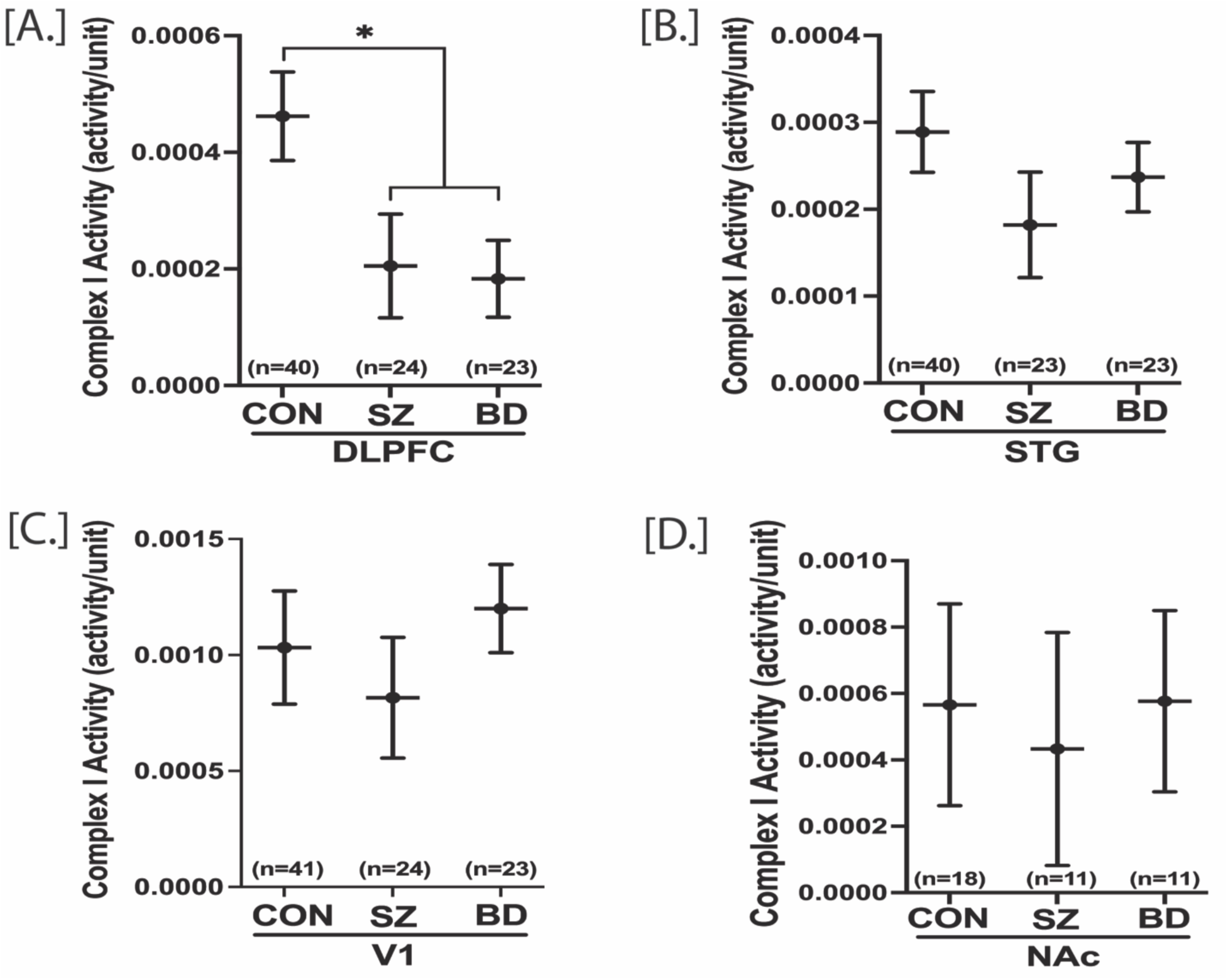
Complex I activity in postmortem DLPFC, STG, V1 and NAc of control (CON), schizophrenia (SZ) and bipolar disorders (BD) groups. Mixed model analysis was performed for statistical significance with adjustment of age and sex. DLPFC Complex I activity was significantly lower in both SZ and BD compared to control group [A]. There were no significant groups differences in STG, V1 and NAc [B, C, D]. All data are represented as least square mean (LSM) ± Std. error. p<0.05 is denoted by *, otherwise p>0.05.

### Overall Mitochondria Number, Synapse Number and Synaptic Mitochondria Number in SZ and BD

The synaptic strength and synaptic activity are dependent upon ATP production by mitochondria. Thus, mitochondria play a pivotal role in synaptic neurotransmission. Among four brain regions studied here, DLPFC has been the most affected area in terms of mtDNA CN, mtDNA deletions and Complex I activity. Therefore, we investigated if there is any change in overall mitochondria number, synapse number and synaptic mitochondria number in DLPFC of SZ and BD subjects. The overall mitochondria puncta number, synapse puncta number and synaptic mitochondria puncta colocalization were determined through immunostaining of mitochondria surface marker TOM40, synaptic marker PSD95 and colocalization of TOM40+PSD95, respectively. The mixed model analysis was conducted with adjustment of subjects’ age, pH and sex. For overall synapse number (# of PSD95 puncta), the main effect of group was significant. The number of synaptic marker (number of PSD95 puncta) was significantly lower in DLPFC of both SZ and BD (p=0.0008) subjects compared to control subjects (Figure 5B). For overall mitochondria number (# of TOM40), there was not significant main effect of group. The number of TOM40 was not significantly different in SZ (p=0.388) and BD (p=0.065) groups compared to CON group. For the synaptic mitochondria number (colocalized TOM40 with PSD95), the main effect of group was not significant. But very interestingly, the sex x group interaction was statistically significant. Both the female SZ (p=0.002) and female BD (p=0.266) subject had significantly lower colocalized TOM40 with PSD95 compared to CON female subjects. The male subjects with SZ and BD did not differ from male control subjects colocalized mitochondria (p = 0.85, p = 0.09, respectively) (Figure 5C-E) (Table ST6). The age did not have any significant effect on the number of PSD95, TOM40 and colocalized PSD95 with TOM40 (Table ST13). Similarly, the number of PSD95, TOM40 and colocalized PSD95 with TOM40 was not dependent on medications (Table ST14).

**Figure 5:**
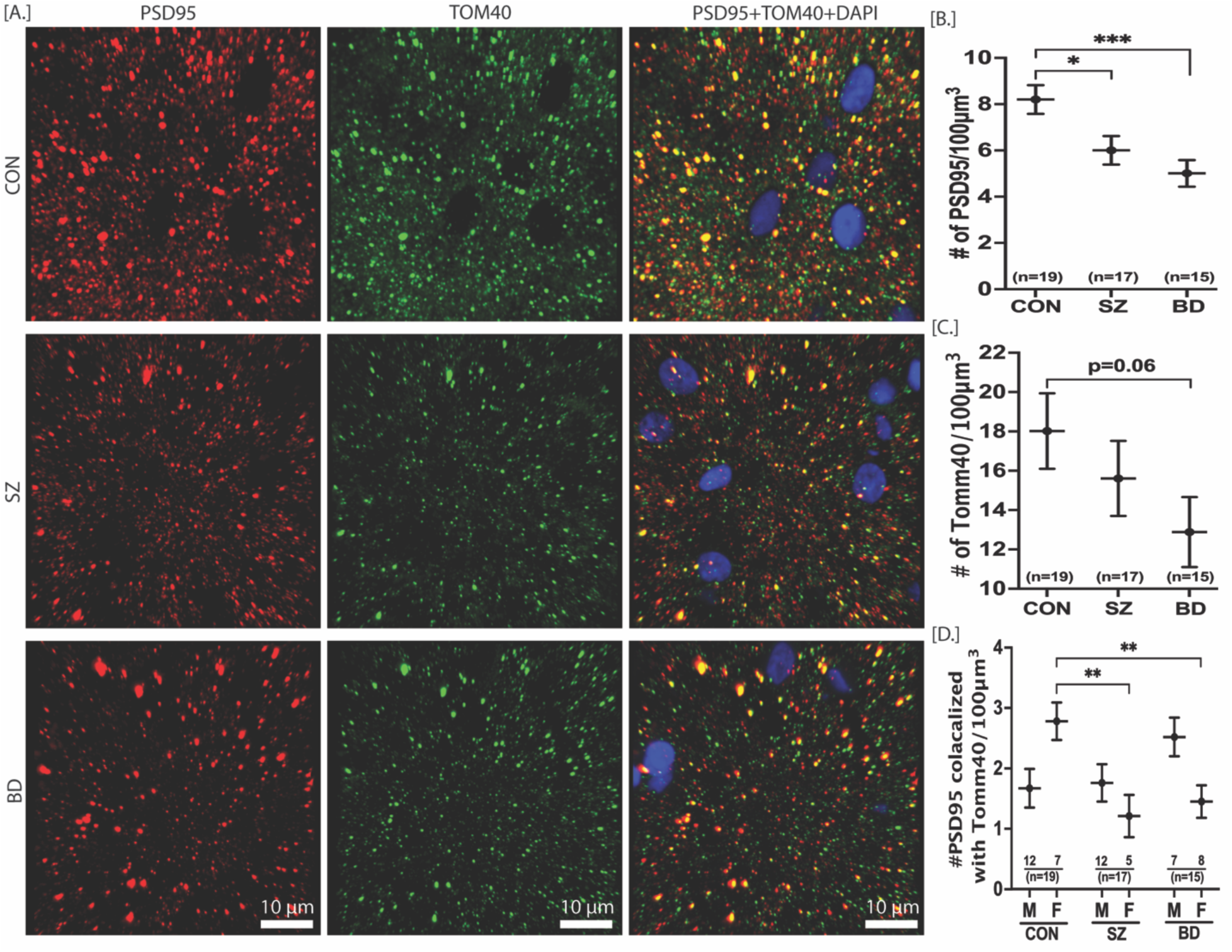
Overall mitochondria number, synapse number and synaptic mitochondria number in DLPFC of CON, SZ and BD subjects. The representative images for PSD95, TOM40 and PSD95 colocalized with TOM40 are depicted in [A]. Mixed model analysis was performed for statistical significance with adjustment of age and sex. The number of PSD95 was significantly lower in both SZ and BD compared to CON group [B]. The overall number of TOM40 was not significantly different in SZ and BD compared to CON, although there was a decrease trend in BD [C]. The sex x group effect was significant for the PSD95 colocalized with TOM40. Both the female SZ and female BD subjects had significantly lower PSD95 colocalized with TOM40 compared to CON females [D]. All data are represented as least square mean (LSM) ± Std. error. p<0.05, p<0.01 and p<0.001 are denoted by *, ** and ***, respectively, otherwise p>0.05. CON=control (n=19), SZ= schizophrenia (n=17), BD=bipolar disorder (n=15), Red spots= PSD95, green spots= TOM40, yellow spots= PSD95 colocalized with TOM40, blue=DAPI nuclear stain.

## Discussion

### Regional Alteration of mtDNA Copy Number in SZ and BD

In this manuscript, we report that postmortem brain tissue of schizophrenia patients showed increased mtDNA CN in DLPFC with no alterations of mtDNA CN in STG, V1 and NAc. On the other hand, bipolar patients showed increased mtDNA CN in DLPFC, decreased mtDNA CN in STG and unaltered mtDNA CN in V1 and NAc (Figure 1). These mtDNA CN results suggest that although schizophrenia and bipolar disorder have overlapping effect on mtDNA CN in various brain regions, but it is not the same for all brain regions. The decreased mtDNA CN has also been reported in postmortem hippocampus of BD patients [27]. Interestingly, peripheral mtDNA CN showed marked decrease in both SZ [21,28,29] and BD [20,30–32] while our postmortem DLPFC mtDNA CN showed marked increase. This discrepancy might be because the brain tissue has higher bioenergetics profile compared to peripheral tissue. Aging is one of the confounding factors affecting peripheral mtDNA CN. Peripheral mtDNA CN has been shown to decrease with aging [33]. Here, we have not found any effect of age on mtDNA CN in all four brain regions, although a trend for decrease with age was observed and warrants further investigation in larger cohorts (Table ST7). These results suggest that aging affect the mtDNA CN parameter in tissue specific manner. In both SZ and BD, the mtDNA copy number changes across brain regions are independent by correlation agreeing other lab reports [34]. The lack of correlation of mtDNA copy number in various brain regions suggests varying energetic demands in each region and perhaps variation in cell types. The mtDNA CN changes in this study were also independent of psychotropic medications or drugs in the postmortem brain (Table ST8).

### The mtDNA common deletion is increased in DLPFC of BD while total mtDNA deletions are increased in DLPFC of both SZ and BD subjects

The mtDNA common deletion (CD) is a 4977 bp deletion which occur due to various events including mtDNA replication/repair, oxidative stress, exposure to ionizing radiation, and mitochondria disease. The mtDNA common deletion can also accumulate as a somatic heteroplasmic deletion with age (positively correlates with age). The age-related accumulation rate of mtDNA CD is tissue specific and more pronounced in tissues with higher energy demands. Here we sought to determine if there is any schizophrenia- or bipolar disorder-specific alteration of mtDNA CD, and at the same time, if there was any region-specific effect. We report here that the mtDNA CD remain unaltered in all four brain regions (DLPFC, STG, V1 and NAc) of SZ subjects. On the other hand, mtDNA CD is increased in DLPFC and unaltered in STG, V1 and NAc of BD subjects (Figure. 2). Previously, we showed that there was a global decrease of mtDNA CD in SZ subjects across 10 brain regions [12]. In contrast to the global effect previously reported in 10 brain regions in SZ, the mtDNA CD was analyzed in four brain regions reducing the statistical power of SZ subjects. The unaltered cortical mtDNA CD in SZ mirrors the previously published results from our and other labs [11,25,35]. The increased mtDNA CD in DLPFC of BD subjects is in accordance with previous findings from our lab determined with independent samples [11]. Previously we showed that the mtDNA common deletion was neither the most frequent nor the most abundant deletion in human brain tissues [18]. This has prompted us to explore the burden of total somatic deletions of mtDNA in schizophrenia and bipolar disorders. Here, we report that the total deletion burden (residuals of deletion per 10K coverage) is increased in DLPFC of both SZ and BD subjects (Figure 3). The residuals of deletion per 10K was not significantly altered in STG, V1 and NAc of both SZ and BD subjects. This is also consistent with our previous observation that the cumulative deletion burden of the DLPFC, relative to the same subject’s anterior cingulate cortex (ACC), was increased in a combined group of SZ and BD subjects when compared to subjects with no psychiatric disease, depression or alcohol dependency [18]. As expected, the deletions per 10K coverage was independent of age and medications in all four brain regions (Table ST10, Figure SF4).

### Decreased Complex I activity in DLPFC of both SZ and BD

We have used a dual method to measure Complex I activity that takes into account the amount of Complex I protein present, then equalizes that amount in the second step of the assay and measures the conversion of NAD+ to NADH. This measure is analogous to a specific activity per unit of enzyme. We did not specifically test for protein abundance deficits, instead choosing to carefully normalize values of Complex I protein for activity measures. The mtDNA CD removes 7 genes encoding 4 Complex I subunits. Thus, we expected decreased Complex I activity in DLPFC of BD subjects since mtDNA CD was increased in DLPFC of BD subjects (Figure. 2). As expected, the Complex I activity was significantly decreased in DLPFC with no alteration in STG, V1 and NAc of BD subjects (Figure. 4). Although the mtDNA CD was unaltered in DLPFC of SZ subjects (Fig. 2), we found that the Complex I activity was also decreased in DLPFC with no alteration in STG, V1 and NAc of SZ subjects (Figure. 4). One possible reason might be due to the increased rate of total mtDNA deletion in DLPFC of SZ subjects as evident by increased residuals of deletion per 10K in DLPFC of SZ subjects. Our decreased DLPFC Complex I activity markedly contrasts with the peripheral platelet Complex I activity of SZ (active psychosis) and BD subjects where the Complex I activity is significantly increased [36–38]. Thus, the peripheral and brain Complex I activity might get affected by different molecular mechanism in blood and brain. The recent meta-analysis [26] showed that the SZ and BD had moderate (statistically insignificant) effect on Complex I activity. This discrepancy might be attributed to the regional heterogeneity of alteration in Complex I activity by SZ and BD. Our postmortem Complex I findings are in line with previously published reports showing decreased Complex I activity in DLPFC and temporal cortex of SZ and BD subjects [13,14,39,40].

### Loss of synapses and sex-specific loss of synaptic mitochondria in both SZ and BD subjects

The energy produced by the mitochondria is utilized in neuronal communication such as development of cytoskeleton for presynapse development, synaptic vesicle transport and generation of synaptic membrane potentials. Mitochondria also plays a crucial role in synaptic neurotransmission through intracellular Ca^2+^ buffering. Thus, the number of mitochondria remain higher in high energy demanding sites such as axon terminals and postsynaptic areas compared to other parts of the neurons [41]. Fewer mitochondria number and/or dysfunctional mitochondria would lead to fewer synapse number or aberrant synaptic communication. In this manuscript, we identified if there is any alteration of the overall mitochondria number, synapse number and synaptic mitochondria number in SZ and BD after adjustment of age and sex. We report here that the overall synapse number are significantly decreased in DLPFC of both SZ and BD subjects. This is an important cellular hallmark in understanding the pathophysiology of SZ and BD. This finding mimics the recent studies showing the decrease of excitatory synapses or postsynaptic elements in prefrontal cortex and anterior cingulate cortex of schizophrenia subjects [42–44]. In contrast to the synapse number, the overall mitochondria number (as measured by IHC that was not restricted to excitatory synapses) was not significantly altered in SZ and BD compared to controls. Decreased mitochondria number has been reported in the axon terminals and layer 3 excitatory synapses of anterior cingulate cortex [40,44,45], and in the oligodendrocytes of prefrontal cortex of SZ subjects [46, 47]. This discrepancy might be attributed to the fact that our calculated overall mitochondria belong to all types of cells and all parts of cells compared to high energy demanding sites such as axon terminals or synapses. To calculate the synaptic mitochondria number, we measured the colocalized spots for mitochondria marker TOM40 with synapse marker PSD95. Interestingly, we have found that synaptic mitochondria number are decreased in both female SZ and female BD subject compared to female controls.

## Conclusion

Among the four brain regions, the DLPFC remains the most affected brain region in terms of mitochondrial parameters studied in this manuscript. The measures of mitochondria unique deletions and copy number are both increased, while Complex I activity is decreased in the DLPFC of both schizophrenia and bipolar subjects compared to controls. Decreased excitatory synapses in schizophrenia and bipolar disorder are relevant to these findings. These indicators of mitochondria dysfunction require further investigations into cell specificity of the findings and potential relationship to genetic influences to understand the pathophysiology of schizophrenia and bipolar disorders.

## Data Availability

All data produced in the present study are available upon reasonable request to the authors

## Acknowledgements

We would like to thank donors’ family for donating the postmortem brain tissues.

## Authors Contributions

Designed, supervised study, and wrote manuscript (MPV). Conducted laboratory experiments (SCD, LM, BR, BH). Data analysis (SCD, BH, MPV, AAO). Drafted and edited manuscript (all).

## Supplementary Information

**Table ST1:** Demographics of subjects

| **A. Demographics of three groups analyzed for Complex I activity, mtDNA common deletion level, mtDNA copy number, and mtDNA total deletion in DLPFC, STG and V1 cortex.** |  |  |  |  |
| --- | --- | --- | --- | --- |
| **Sex** |  | **BD** | **CON** | **SZ** |
| Female (N) |  | 13 | 6 | 3 |
| Male (N) |  | 13 | 35 | 24 |
|  | F : M | 1:01 | 1:06 | 1:8 # |
| pH | Mean | 6.68* | 6.86 | 6.64* |
|  | Std Dev | 0.15 | 0.22 | 0.2 |
| Age | Mean | 46.0* | 57.9 | 41.2* |
|  | Std Dev | 12.5 | 12.7 | 10.1 |
| PMI | Mean | 20.5 | 18.8 | 21 |
|  | Std Dev | 6.61 | 7.12 | 7.39 |
| # of Meds | Mean | 3.7* | 0.9 | 2.1* |
|  | Std Dev | 3.2 | 1.7 | 1.7 |
|  | Min | 0 | 0 | 0 |
|  | Max | 10 | 10 | 5 |
| # Number of male and female subjects is different between groups |  |  |  |  |
| * Significantly different from control group (p < 0.05). |  |  |  |  |
| **C. Demographics of three groups analyzed for colocalization of TOMM40 and PSD95 in DLPFC.** |  |  |  |  |
| **Sex** |  | **BD** | **CON** | **SZ** |
| Female (N) |  | 8 | 7 | 5 |
| Male (N) |  | 7 | 12 | 12 |
|  | F : M<sup>#</sup> | 1:0.88 | 1:1.7 | 1:2.4 |
| pH | Mean | 6.62** | 6.85 | 6.67* |
|  | Std Dev | 0.12 | 0.23 | 0.21 |
| Age | Mean | 43.92*** | 64.1 | 43.5*** |
|  | Std Dev | 11.92 | 14.32 | 11.97 |
| PMI | Mean | 19.69 | 18.98 | 22.59 |
|  | Std Dev | 6.15 | 6.51 | 6.11 |
| # Chi-square statistic 2.99 (N = 51, p-value = 0.22) for sex distribution by group. |  |  |  |  |
| * p < 0.05, ** p < 0.005, *** p < 0.0005, compared to control. |  |  |  |  |
| **B. Subset of samples depicted in (A) analyzed for complex I activity, mtDNA common deletion level, mtDNA copy number, and mtDNA total deletion in NAc.** |  |  |  |  |
| **Gender** |  | **BD** | **CON** | **SZ** |
| Female (N) |  | 7 | 3 | 1 |
| Male (N) |  | 9 | 18 | 10 |
| All |  | 16 | 21 | 11 |
|  | **F:M** | **0.8:1** | **1:06** | **1:10** |
| Age | Mean | 43.81*** | 60.76 | 43.00*** |
| Age | Std Dev | 10.51 | 13.91 | 11.8 |
| pH | Mean | 6.71* | 6.87 | 6.63* |
| pH | Std Dev | 0.14 | 0.28 | 0.28 |
| PMI | Mean | 20.82 | 17.72 | 20.96 |
| PMI | Std Dev | 6.84 | 8.53 | 6.94 |
| *** p < 0.0005, ** p < 0.005, * p < 0.05 |  |  |  |  |

**Table ST2:** mtDNA copy number in four brain regions among control, schizophrenia and bipolar disorders.

| DLPFC mtDNA Copy Number--qPCR |  |  |  |  | V1 mtDNA Copy Number--ddPCR |  |  |  |
| --- | --- | --- | --- | --- | --- | --- | --- | --- |
|  | Least Square Mean | Std Error | p-Value |  |  | Least Square Mean | Std Error | p-Value |
| CON | 1394.3 | 79 |  |  | CON | 1697.8 | 127.9 |  |
| SZ | 1799.5 | 110.4 | 0.003 |  | SZ | 1541.7 | 167.1 | 0.46 |
| BD | 1702.9 | 91.9 | 0.011 |  | BD | 1684.4 | 140.5 | 0.94 |
| STG mtDNA Copy Number--qPCR |  |  |  |  | NAc mtDNA Copy Number--qPCR |  |  |  |
|  | Least Square Mean | Std Error | p-Value |  |  | Least Square Mean | Std Error | p-Value |
| CON | 1661.9 | 79 |  |  | CON | 6016.3 | 1008.2 |  |
| SZ | 1492.1 | 106.4 | 0.202 |  | SZ | 6369.1 | 1265.5 | 0.82 |
| BD | 1406.2 | 91.9 | 0.036 |  | BD | 7700.4 | 722.1 | 0.17 |

**Table ST3:** mtDNA common deletion in four brain regions among control, schizophrenia and bipolar disorders.

| DLPFC mtDNA Common Deletion--qPCR |  |  |  |  | V1 mtDNA Common Deletion--qPCR |  |  |  |
| --- | --- | --- | --- | --- | --- | --- | --- | --- |
|  | Least Square Mean | Std Error | p-Value |  |  | Least Square Mean | Std Error | p-Value |
| CON | 0.082 | 0.01 |  |  | CON | 0.021 | 0.01 |  |
| SZ | 0.065 | 0.015 | 0.36 |  | SZ | 0.037 | 0.015 | 0.39 |
| BD | 0.128 | 0.012 | 0.004 |  | BD | 0.04 | 0.013 | 0.24 |
| STG mtDNA Common Deletion--qPCR |  |  |  |  | NAc mtDNA Common Deletion--qPCR |  |  |  |
|  | Least Square Mean | Std Error | p-Value |  |  | Least Square Mean | Std Error | p-Value |
| CON | 0.006 | 0.01 |  |  | CON | 0.074 | 0.013 |  |
| SZ | 0.007 | 0.014 | 0.96 |  | SZ | 0.066 | 0.017 | 0.73 |
| BD | 0.011 | 0.012 | 0.72 |  | BD | 0.06 | 0.014 | 0.54 |

**Table ST4:**
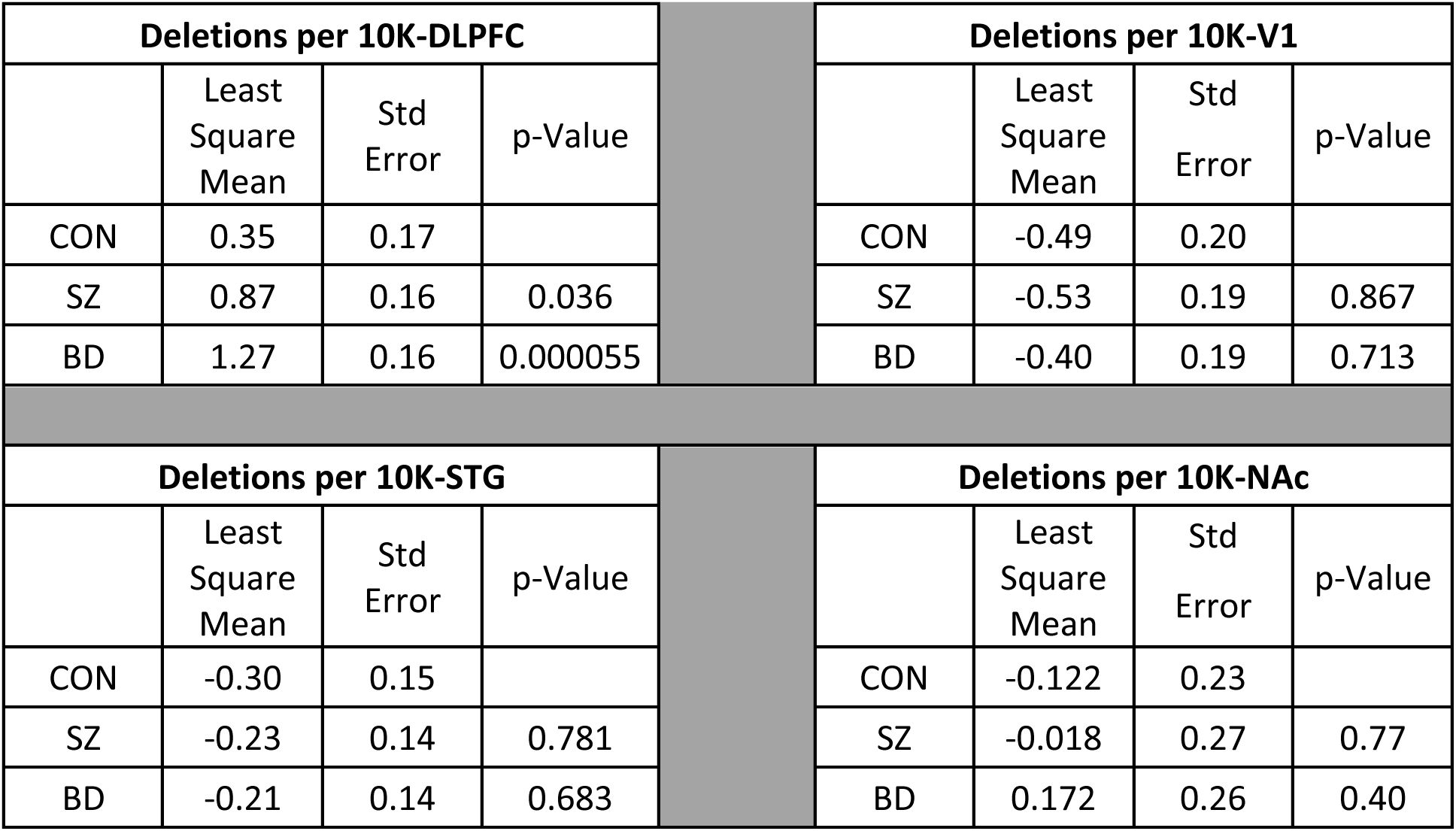
mtDNA deletions per 10K in four brain regions among control, schizophrenia and bipolar disorders.

| Deletions per 10K-DLPFC |  |  |  |  | Deletions per 10K-V1 |  |  |  |
| --- | --- | --- | --- | --- | --- | --- | --- | --- |
|  | Least Square Mean | Std Error | p-Value |  |  | Least Square Mean | Std Error | p-Value |
| CON | 0.35 | 0.17 |  |  | CON | -0.49 | 0.20 |  |
| SZ | 0.87 | 0.16 | 0.036 |  | SZ | -0.53 | 0.19 | 0.867 |
| BD | 1.27 | 0.16 | 0.000055 |  | BD | -0.40 | 0.19 | 0.713 |
| Deletions per 10K-STG |  |  |  |  | Deletions per 10K-NAc |  |  |  |
|  | Least Square Mean | Std Error | p-Value |  |  | Least Square Mean | Std Error | p-Value |
| CON | -0.30 | 0.15 |  |  | CON | -0.122 | 0.23 |  |
| SZ | -0.23 | 0.14 | 0.781 |  | SZ | -0.018 | 0.27 | 0.77 |
| BD | -0.21 | 0.14 | 0.683 |  | BD | 0.172 | 0.26 | 0.40 |

**Table ST5:** Complex I activity in four brain regions among control, schizophrenia and bipolar disorders.

| Complex I activity-DLPFC |  |  |  |  | Complex I activity-V1 |  |  |  |
| --- | --- | --- | --- | --- | --- | --- | --- | --- |
|  | Least Square Mean | Std Error | p-Value |  |  | Least Square Mean | Std Error | p-Value |
| CON | 0.00046 | 0.00007 |  |  | CON | 0.0010 | 0.0002 |  |
| SZ | 0.00020 | 0.00008 | 0.03 |  | SZ | 0.0008 | 0.0003 | 0.53 |
| BD | 0.00018 | 0.00006 | 0.01 |  | BD | 0.0012 | 0.0002 | 0.61 |
| Complex I activity-STG |  |  |  |  | Complex I activity-NAc |  |  |  |
|  | Least Square Mean | Std Error | p-Value |  |  | Least Square Mean | Std Error | p-Value |
| CON | 0.00029 | 0.00005 |  |  | CON | 0.0006 | 0.0003 |  |
| SZ | 0.00018 | 0.00006 | 0.16 |  | SZ | 0.0004 | 0.0004 | 0.77 |
| BD | 0.00023 | 0.00004 | 0.43 |  | BD | 0.0006 | 0.0003 | 0.98 |

**Table ST6:**
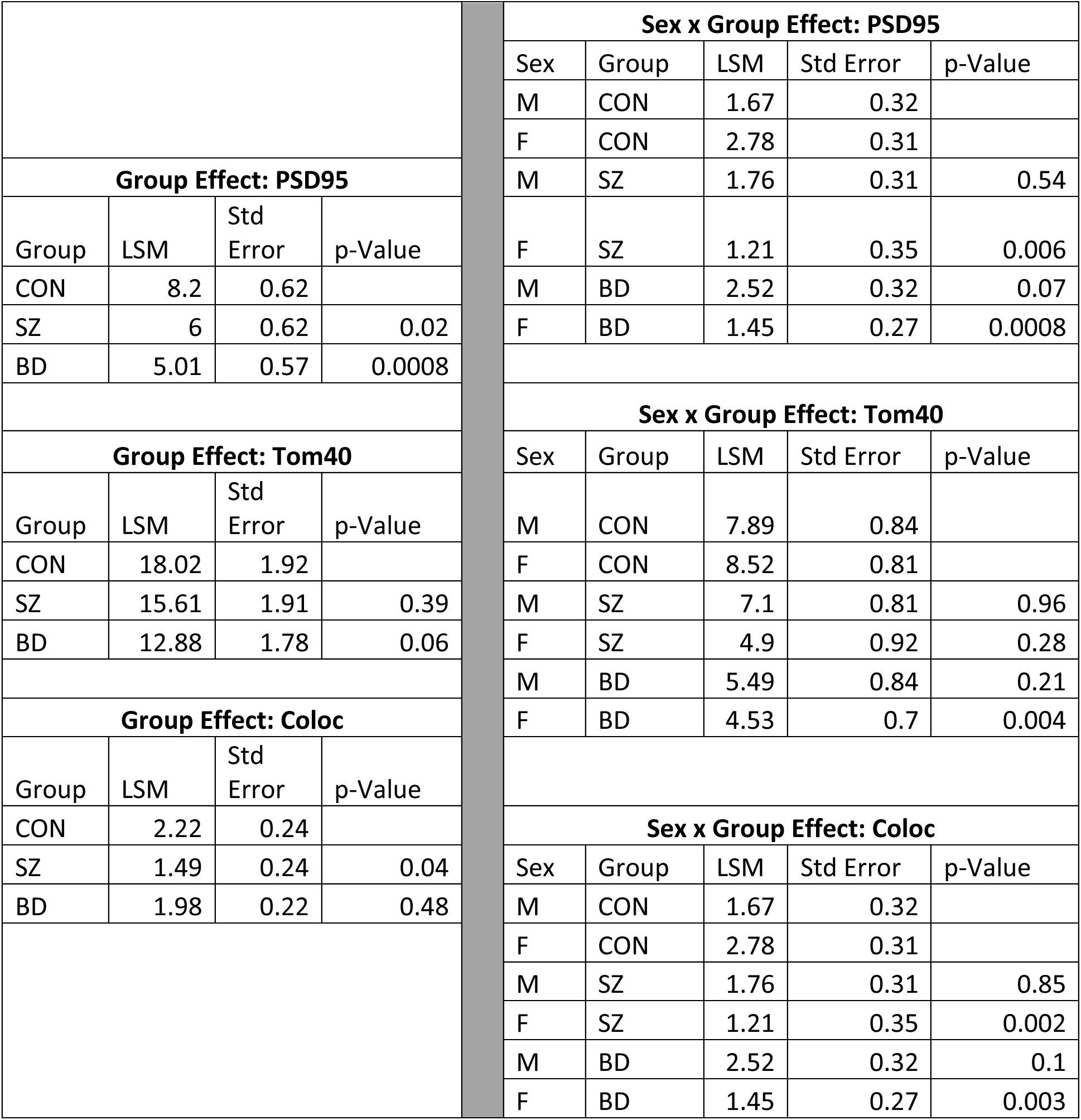
Number of overall mitochondria number, synapse number and synaptic mitochondria number in DLPFC of CON, SZ and BD subjects.

**Table ST7:** Correlation of age on mtDNA copy number in DLPFC, STG, V1 and NAc.

|  | **DLPFC** | **STG** | **V1** |
| --- | --- | --- | --- |
| Pearson r | -0.18009 | -0.20203 | -0.00118 |
| p-Value | 0.087761 | 0.050887 | 0.992 |
| df | 93 | 94 | 86 |

**Table ST8:** Effect of Medications on mtDNA copy number in DLPFC, STG, V1 and NAc. 1 = Pooled BD and SZ subjects positive for neuroactive medications by toxicology screen 2= Pooled BD and SZ subjects negative for neuroactive medications by toxicology screen C = controls

| **Region** | **1** | **2** | **controls** | **t-test (p-value)** |  | **Region** | **1** | **2** | **controls** | **t-test (p-value)** |
| --- | --- | --- | --- | --- | --- | --- | --- | --- | --- | --- |
| **DLPFC** |  |  |  | **1,2** |  | **STG** |  |  |  | **1,2** |
| Mean | 1691.461 | 1992.11 | 1413.26 | 0.05868 |  | Mean | 1495.799 | 1430.379 | 1538.43 | 0.521886 |
| Stdev | 457.7294 | 624.761 | 489.919 |  |  | Stdev | 326.4067 | 356.95 | 421.657 |  |
| Fold Change | 1.196852 | 1.40958 |  | 1, C |  | Fold Change | 0.97229 | 0.929767 |  | 1, C |
|  |  |  |  | 0.012616 |  |  |  |  |  | 0.621633 |
|  |  |  |  | 2,C |  |  |  |  |  | 2,C |
|  |  |  |  | 0.000461 |  |  |  |  |  | 0.363372 |
| **Region** | **1** | **2** | **controls** | **t-test (p-value)** |  | **Region** | **1** | **2** | **controls** | **t-test (p-value)** |
| **V1** |  |  |  | **1,2** |  | **NAc** |  |  |  | **1,2** |
| Mean | 1611.813 | 1567.75 | 1665.46 | 0.832819 |  | Mean | 8062.819 | 8644.295 | 7099.24 | 0.642232 |
| Stdev | 737.8149 | 471.97 | 605.694 |  |  | Stdev | 2156.662 | 4202.303 | 2639.04 |  |
| Fold Change | 0.967787 | 0.94133 |  | 1, C |  | Fold Change | 1.13573 | 1.217637 |  | 1, C |
|  | 1/c | 2/c |  | 0.739918 |  |  | 1/c | 2/c |  | 0.273168 |
|  |  |  |  | 2,C |  |  |  |  |  | 2,C |
|  |  |  |  | 0.579958 |  |  |  |  |  | 0.230739 |

**Table ST9:** Inter-region correlation and age-effect on mtDNA common deletion in DLPFC, STG, V1 and NAc.

| **Correlation** | **CD%-STG** | **CD%-DLPFC** | **CD%-NAc** | **CD %-V1** | **Age** |
| --- | --- | --- | --- | --- | --- |
| **CD%-STG** | 1 | 0.758212 | 0.462624 | 0.821403 | 0.273021 |
| **CD%-DLPFC** | 0.758212 | 1 | 0.531127 | 0.718525 | 0.344293 |
| **CD%-NAc** | 0.462624 | 0.531127 | 1 | 0.369471 | 0.625985 |
| **CD %-V1** | 0.821403 | 0.718525 | 0.369471 | 1 | 0.22768 |
| **Age** | 0.273021 | 0.344293 | 0.625985 | 0.22768 | 1 |
| **p-Value** | **CD%-STG** | **CD%-DLPFC** | **CD%-NAc** | **CD %-V1** | **Age** |
| **CD%-STG** | 0 | 3.25E-18 | 0.00106 | 1.11E-22 | 0.008101 |
| **CD%-DLPFC** | 3.25E-18 | 0 | 0.000123 | 3.24E-15 | 0.000833 |
| **CD%-NAc** | 0.00106 | 0.000123 | 0 | 0.010591 | 2.53E-06 |
| **CD %-V1** | 1.11E-22 | 3.24E-15 | 0.010591 | 0 | 0.032892 |
| **Age** | 0.008101 | 0.000833 | 2.53E-06 | 0.032892 | 0 |

**Table ST10:** Inter-region correlation and age-effect on deletions per 10K across DLPFC, STG, V1 and NAc.

| **Correlation** | **Age** | **DLPFC** | **NAc** | **STG** | **V1** |
| --- | --- | --- | --- | --- | --- |
| **Age** | 1 | 0.078035 | -0.00125 | -0.06097 | 0.008678 |
| **DLPFC** | 0.078035 | 1 | -0.01485 | 0.050401 | 0.146984 |
| **NACC** | -0.00125 | -0.01485 | 1 | -0.1166 | -0.29514 |
| **STG** | -0.06097 | 0.050401 | -0.1166 | 1 | 0.00112 |
| **V1** | 0.008678 | 0.146984 | -0.29514 | 0.00112 | 1 |
| **p-Value** | **Age** | **DLPFC** | **NACC** | **STG** | **V1** |
| **Age** | 0 | 0.454702 | 0.993438 | 0.563717 | 0.93457 |
| **DLPFC** | 0.454702 | 0 | 0.921972 | 0.633274 | 0.162066 |
| **NAc** | 0.993438 | 0.921972 | 0 | 0.440285 | 0.046454 |
| **STG** | 0.563717 | 0.633274 | 0.440285 | 0 | 0.991544 |
| **V1** | 0.93457 | 0.162066 | **0.046454** | 0.991544 | 0 |

**Table ST11:** Inter-region correlation and age-effect on Complex I activity across DLPFC, STG, V1 and NAc.

| Multivariate Correlation | Complex I activity STG | Complex I Activity DLPFC | Complex I activity V1 | Complex I Activity NAc | Age |
| --- | --- | --- | --- | --- | --- |
| Complex I activity-STG | **1.0000** | 0.1768 | 0.0611 | 0.0417 | 0.2694 |
| Complex I activity-DLPFC | 0.1768 | **1.0000** | -0.0927 | 0.0328 | 0.1513 |
| Complex I activity-V1 | 0.0611 | -0.0927 | **1.0000** | 0.1305 | -0.0171 |
| Complex I activity-NAc | 0.0417 | 0.0328 | 0.1305 | **1.0000** | -0.1775 |
| Age | 0.2694 | 0.1513 | -0.0171 | -0.1775 | **1.0000** |

| Correlation Probability | Complex I activity STG | Complex I Activity DLPFC | Complex I activity V1 | Complex I Activity NAc | Age |
| --- | --- | --- | --- | --- | --- |
| Complex I activity-STG | **<.0001** | 0.1122 | 0.5857 | 0.7856 | **0.0144** |
| Complex I activity-DLPFC | 0.1122 | **<.0001** | 0.3988 | 0.8306 | 0.1668 |
| Complex I activity-V1 | 0.5857 | 0.3988 | **<.0001** | 0.3930 | 0.8705 |
| Complex I activity-NAc | 0.7856 | 0.8306 | 0.3930 | **<.0001** | 0.2435 |
| Age | **0.0144** | 0.1668 | 0.8705 | 0.2435 | **<.0001** |

**Table ST12:** Effect of Medication/Drugs on complex I activity in DLPFC, STG, V1 and NAc.

| Multivariate Correlation | Complex I activity STG | Complex I Activity DLPFC | Neuroactive Drugs | Complex I activity V1 | Complex I Activity NAc |
| --- | --- | --- | --- | --- | --- |
| Complex I activity-STG | **<.0001** | 0.1093 | 0.4905 | 0.3879 | 0.5834 |
| Complex I activity-DLPFC | 0.1093 | **<.0001** | **0.0073** | 0.3890 | 0.5848 |
| Neuroactive Drugs | 0.4905 | **0.0073** | **<.0001** | 0.1461 | 0.8634 |
| Complex I activity-V1 | 0.3879 | 0.3890 | 0.1461 | **<.0001** | 0.5101 |
| Complex I activity-NAc | 0.5834 | 0.5848 | 0.8634 | 0.5101 | **<.0001** |

**Table ST13:** Effect of Age on number of TOM40, PSD95 and coloc in DLPFC.

| | Age | PSD95/100$\mu\text{m}^3$ | TOM40/100$\mu\text{m}^3$ | PSD95coloc w/ Tomm40 |
| --- | --- | --- | --- | --- |
| Age | **1.0000** | -0.2159 | -0.0603 | -0.1068 |
| PSD95/100$\mu\text{m}^3$ | -0.2159 | **1.0000** | **0.4116** | **0.5132** |
| TOM40/100$\mu\text{m}^3$ | -0.0603 | **0.4116** | **1.0000** | **0.6865** |
| PSD95coloc w/ Tomm40 | -0.1068 | **0.5132** | **0.6865** | **1.0000** |
| Age | **<.0001** | 0.2794 | 0.7653 | 0.5959 |
| PSD95/100$\mu\text{m}^3$ | 0.2794 | **<.0001** | **0.0329** | **0.0062** |
| TOM40/100$\mu\text{m}^3$ | 0.7653 | **0.0329** | **<.0001** | **<.0001** |
| PSD95coloc w/ Tomm40 | 0.5959 | **0.0062** | **<.0001** | **<.0001** |

**Table ST14:** Effect of Medication/Drugs on number of Tomm40, PSD95 and Coloc in DLPFC.

| | PSD95/100$\mu\text{m}^3$ | TOM40/100$\mu\text{m}^3$ | PSD95coloc w/ Tomm40 | Neuroactive Drugs |
| --- | --- | --- | --- | --- |
| PSD95/100$\mu\text{m}^3$ | **1.0000** | **0.4116** | **0.5132** | -0.3133 |
| TOM40/100$\mu\text{m}^3$ | **0.4116** | **1.0000** | **0.6865** | -0.1724 |
| PSD95coloc w/ Tomm40 | **0.5132** | **0.6865** | **1.0000** | -0.2477 |
| Neuroactive Drugs | -0.3133 | -0.1724 | -0.2477 | **1.0000** |
| PSD95/100$\mu\text{m}^3$ | **<.0001** | **0.0329** | **0.0062** | 0.1116 |
| TOM40/100$\mu\text{m}^3$ | **0.0329** | **<.0001** | **<.0001** | 0.3898 |
| PSD95coloc w/ Tomm40 | **0.0062** | **<.0001** | **<.0001** | 0.2130 |
| Neuroactive Drugs | 0.1116 | 0.3898 | 0.2130 | **<.0001** |

**Figure ST1:**
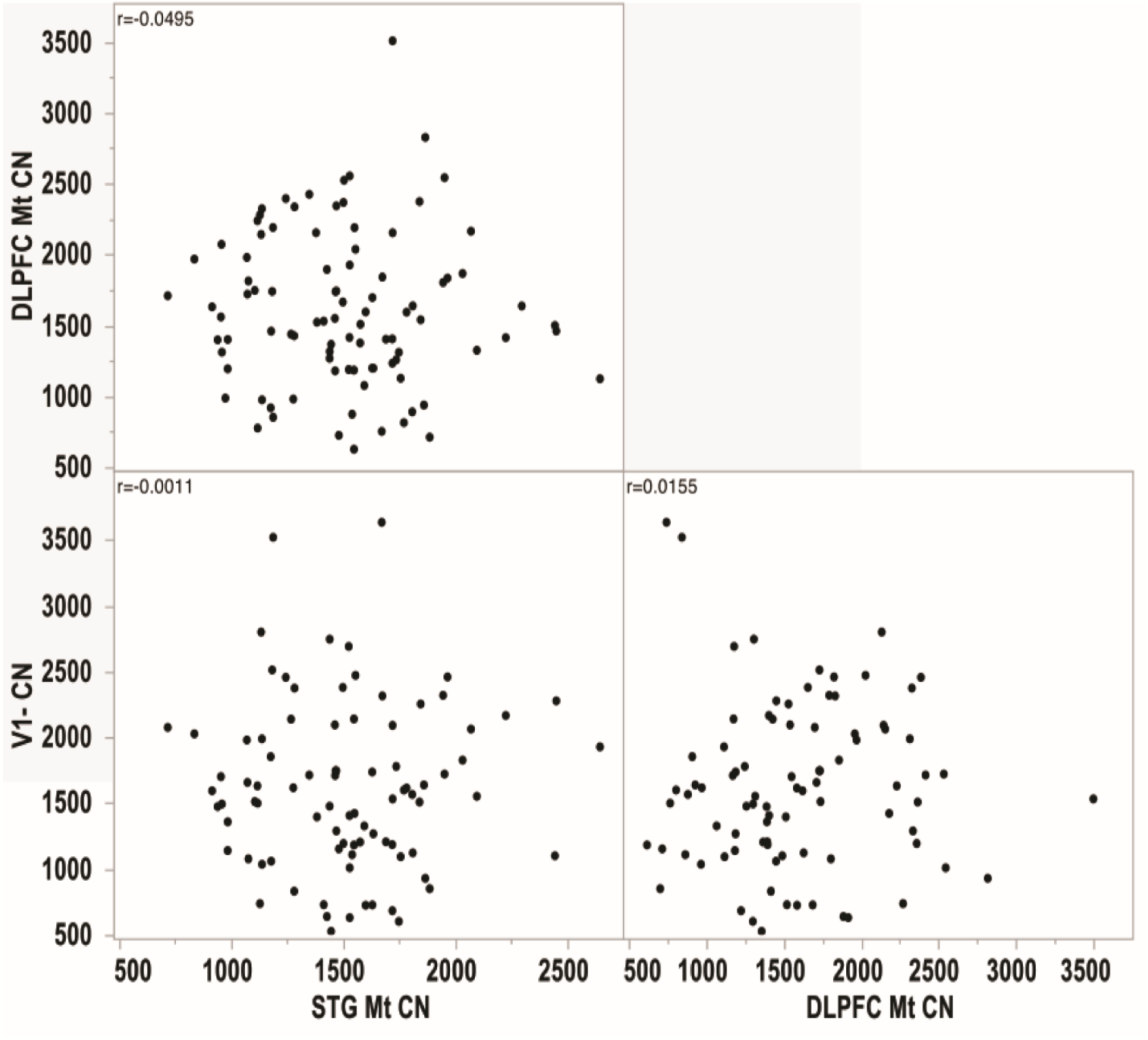
mtDNA copy number correlation across DLPFC, V1 and STG.

**Figure ST2:**
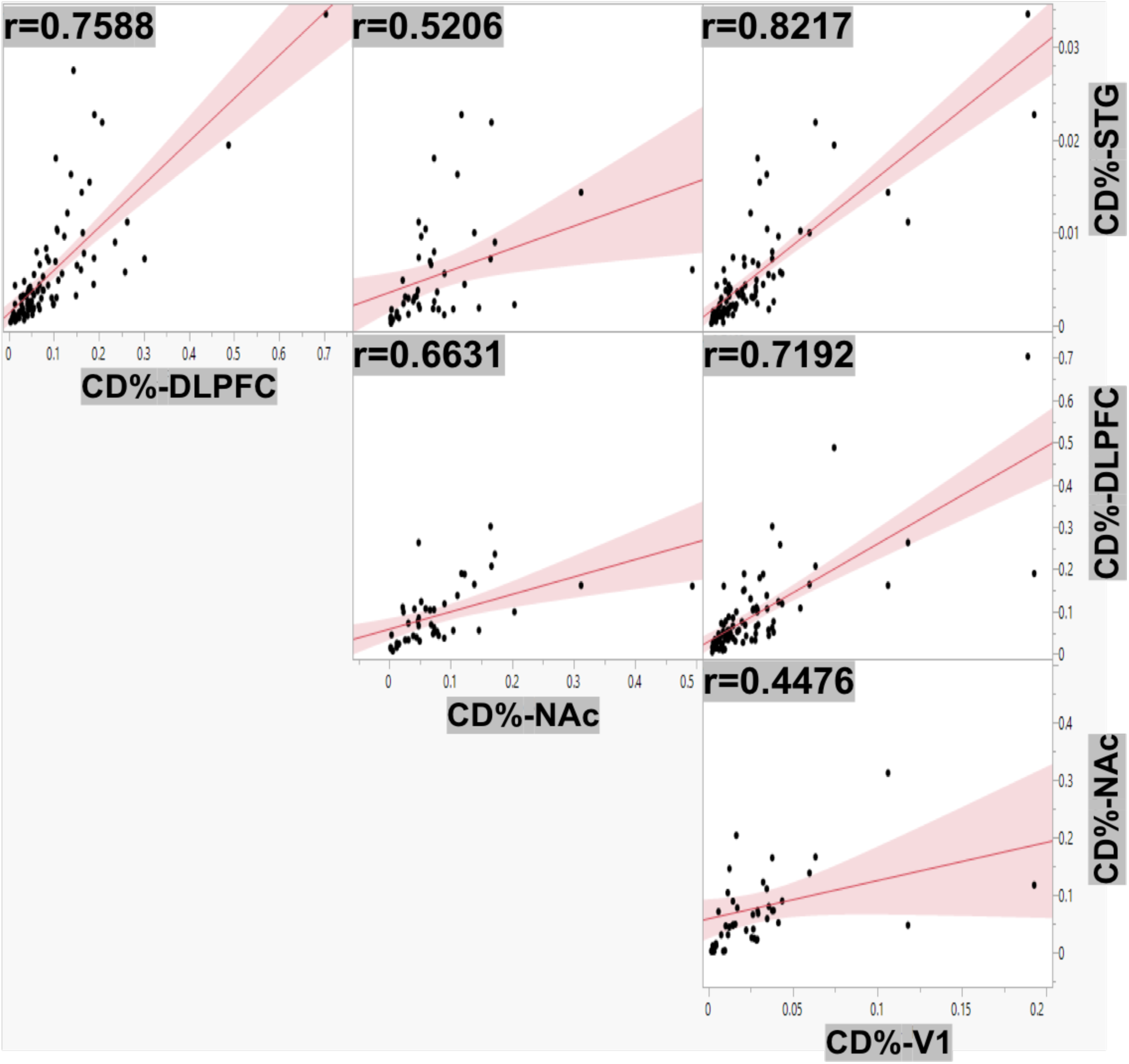
mtDNA common deletion correlation across DLPFC, STG, V1 and NAc.

**Figure ST3:**
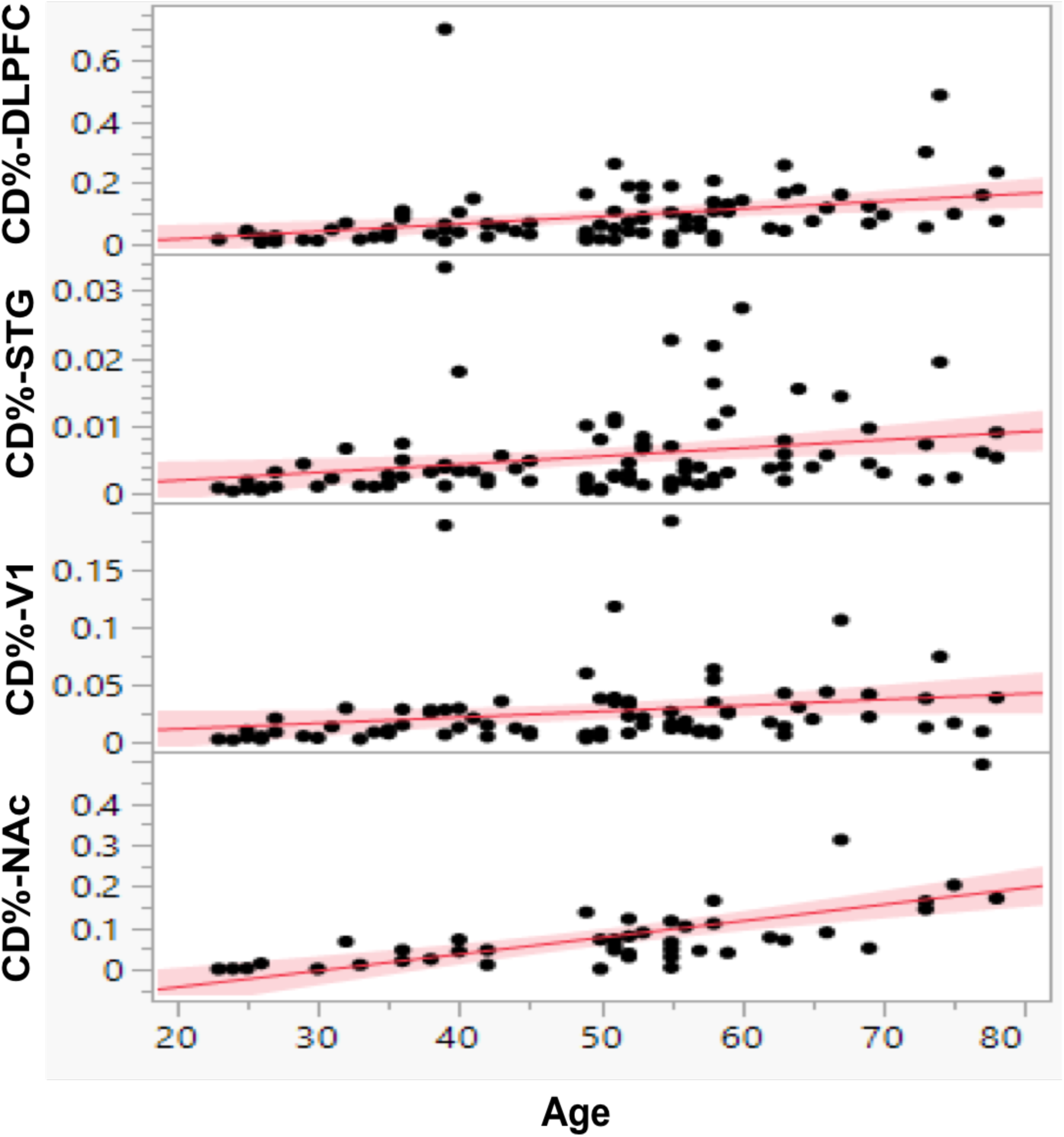
Effect of age on common deletions (%) in DLPFC, STG, V1 and NAc.

**Figure ST4:**
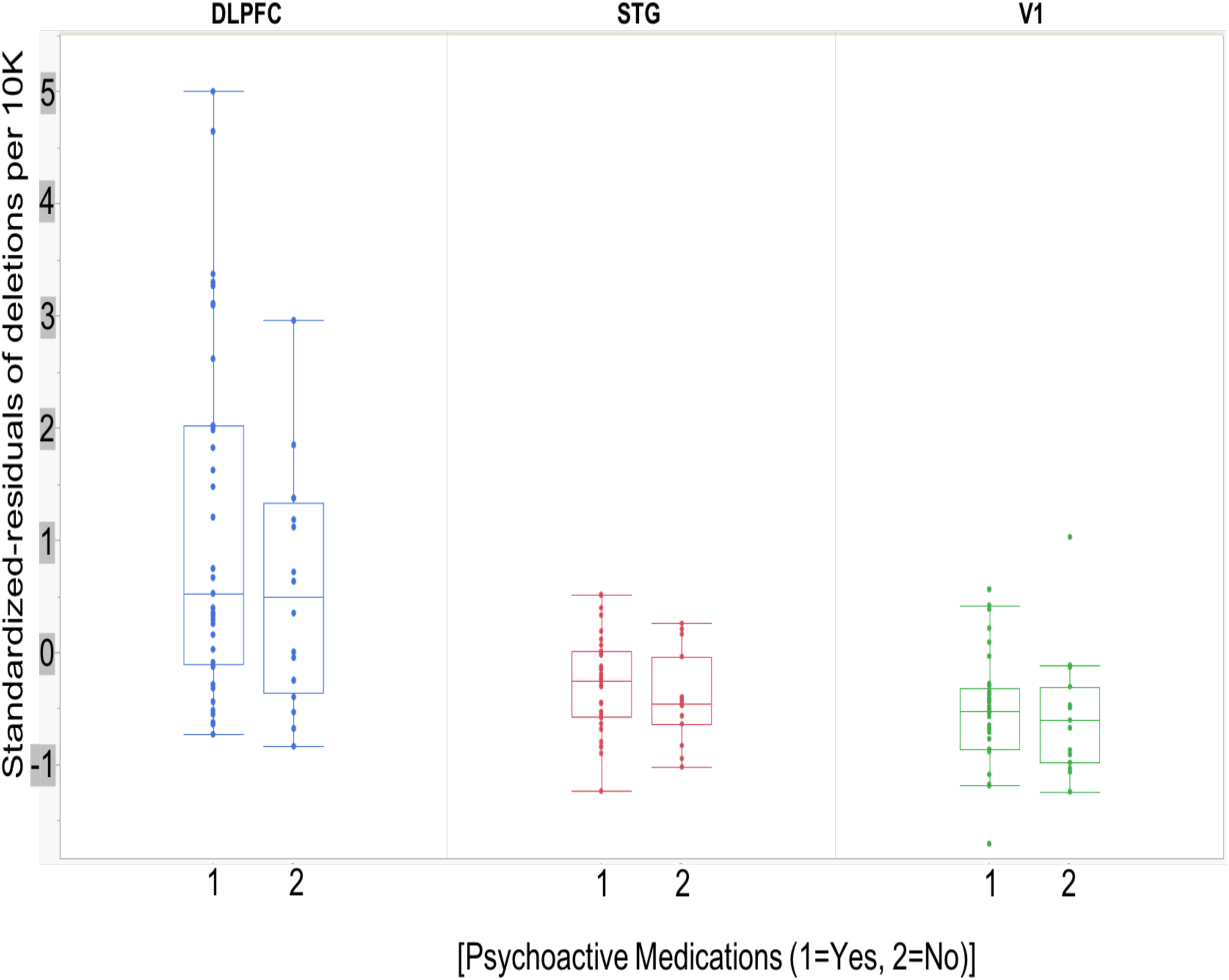
Effect of Medication/Drugs on deletions per 10K in DLPFC, STG and V1.

## Supplementary Materials and Methods

### mtDNA Copy Number and Common Deletion

The genomic DNA was extracted from DLPFC, STG, V1 and NAc samples using a phenol chloroform extraction method. The mtDNA CN and mtDNA CD in DLPFC, STG and NAc samples was determined with quantitative real-time PCR (qPCR), whereas mtDNA CN and mtDNA CD in V1 samples was determined by droplet digital PCR. Quantitative real-time PCR was run using the Applied Biosystems 7900HT real-time instrument in 12-µL reaction volumes using 2 ng gDNA and 2 µl of each standard with published conditions in triplicate. For mtDNA CN, a 200-bp gBlock oligonucleotide (Integrated DNA Technologies, Coralville, IA, USA) representing the human albumin gene (ALB) on chromosome 4 is the reference DNA standard. An 11- point standard curve created with this gBlock has copies ranging from 50 - 50,000 copies per µl of the albumin gene. A 326-bp gBlock Gene Fragment (Integrated DNA Technologies, Coralville, IA) for the mitochondrial wild type sequence (mt13176-13501) is used as a copy number standard for mtDNA. An 11-point standard curve is created with copies ranging from 20,000-10,000,000 per µL of mitochondria wild type product. Mitochondrial DNA CN for the samples is determined using the ratio of mtDNA CN/ALB CN. For the mtDNA CD, a 301-bp gBlock Gene Fragment (Integrated DNA Technologies, Coralville, IA, USA) encompassing the common deletion breakpoints (mt8224–8469 and mt13447–13501) is used as a standard for the deletion. An 11-point standard curve is created with copies ranging from 50-50,000-copies/µL. CD% is calculated using the following equation: CD% = (mtDNA deletion copies)/ (mtDNA wild type copies + deletion copies) × 100.

The following primers were used:

mtDNA CN/CD based on SYBR Green chemistry:

ALB_Primer_F: 5‘-TGCATGAGAAAACGCCAGTA-3’

ALB_Primer_R: 5‘-TCTGCATGGAAGGTGAATGT-3’

mtWT_Primer_F: 5‘-TTACAATCGGCATCAACCAA-3’

mtWT_Primer_R: 5‘-GCTAATGCTAGGCTGCCAAT-3’

mtDel_Primer_F: 5‘-AGGGCCCGTATTTACCCTAT-3’

mtDel_Primer_R: 5‘-GCTAATGCTAGGCTGCCAAT-3’

mtDNA CN and mtDNA CD in V1 samples were determined using the QX200 Droplet Digital PCR System (Bio-Rad, Hercules, CA, USA). For each assay a mixture of both the forward and reverse primers and probes were combined. Stock concentrations of each primer and probe were 100 µM and to make 100 reactions worth of a 20x assay we used 18 µL of forward primer stock at 100 µM, 18 µL of reverse primer stock at 100 µM, 5 ul of probe stock at 100 µM and 59 µL of water. The final 1x concentration was 900 nM for each primer and 250 nM for each probe. Each 20 µL reaction consisted of 10 µL 2x ddPCR Supermix (Bio-Rad, Hercules, CA, USA), 1 µL of the above 20x assay consisting of the primers and probe, and the appropriate amount of gDNA and water up to the final 20 µL volume. To run the mitochondria wildtype assay, 0.01 ng of gDNA were used. To run the ALB and common deletion assays 10 ng of gDNA were used. Samples were placed in the QX200 Droplet Generator to partition each sample into 20,000 nanoliter sized droplets. Droplets were then run on the thermal cycler with the following program: 95 x 10 min, followed by 40 cycles of 94 x 30 sec then 60 x 1 min. A final enzyme deactivation for 98 x 10 min was followed by a hold at 4. Following PCR amplification, the droplets were analyzed in the QX200 Droplet Reader. QuantaSoft Software (Bio-Rad, Hercules, CA, USA) was used to measure the number of positive and negative droplets for each fluorophore.

The following primers and probe Integrated DNA Technologies, Coralville, IA, USA) were used for ddPCR (all as listed 5’-3’):

Primers for ALB: F-TGCATGAGAAAACGCCAGTA, R- TCTGCATGGAAGGTGAATGT Probe for ALB: /5HEX/TGGTGAACA/ZEN/GGCGACCATGCT/3IABkFQ/.

For the mito wildtype (WT) assay, the primers and probe were

Primers for WT: F- GGAAGCCTATTCGCAGGATT, R- CTAGGAAAGTGACAGCGAGG Probe for WT: /56-FAM/CCCCTTCCA/ZEN/AACAACAATCCCC/3IABkFQ/.

For the common deletion (CD) assay the primers and probe were-

Primers for CD: F-AGGGCCCGTATTTACCCTAT, R-GCTAATGCTAGGCTGCCAAT Probe for CD: /5HEX/CCCCTCTAG/ZEN/AGCCCACTGTAAAGC/3IABkFQ/.

### Pooled Effect of Total Deletion of mtDNA by High-resolution Pipeline “Splice-Break”

In this manuscript, we present the results of the number of deletion species per 10K coverage as this showed a significant effect across diagnosis. The number of deletions per 10K was calculated by counting the total number of deletion species (unique set of breakpoints) in a particular sample, then dividing by the benchmark coverage and multiplying by 10000. The Splice-Break pipeline included long-range (LR) PCR amplification of mtDNA, next-generation sequencing (NGS), and MapSplice (v.2.1.18) algorithm (which detect putative mtDNA deletion breakpoint “junctions”). Prior to NGS library preparation, the mtDNA was enriched for each sample using a long-range (LR) PCR utilizing back-to-back primers that hybridize to the control region of the mitochondrial genome. Primer sequences used for most samples were F 5’- CCGCACAAGAGTGCTACTCTCCTC-3’ and R 5’-GATATTGATTTCACGGAGGATGGTG-3’ (Integrated DNA Technologies). In cases where back-to-back primers failed to amplify the WT mitochondrial genome, the following alternate primers were used: 5’- AAGCCTAAATAGCCCACACG-3’ and 5’-GTGGCTTTGGAGTTGCAGTT-3’. For NGS, libraries were prepared using the 384 Seq-Well kit (San Diego, CA). Libraries are sequenced as 150-mer paired-end reads on a patterned flow cell using the Illumina NovaSeq6000 at the UCI Genomics High Throughput Facility. We used our custom bash script Splice-Break.sh (https://github.com/brookehjelm/Splice-Break/) to perform automated filtering, normalization, and annotation of mtDNA deletion breakpoints, and performed manual calculations of the cumulative deletion metrics.

### Complex I Activity (activity/unit of Complex I protein)

Briefly, brain tissue (60–100 mg) was homogenized in PBS at 4°C and centrifuged at 3000 rpm for 5 min. After centrifugation, the first supernatant is collected and measured for protein concentration to ensure adequate protein for both ELISA and activity assays. Equivalent amounts of supernatant protein are extracted (Part 8201086, Abcam, Cambridge, UK), the second supernatants are collected (16000xg, 4°C, 20 min) and stored at –80°C for Complex I ELISA and activity assays. The Complex I ELISA assay (AB178011, Abcam, Cambridge, UK) quantitatively measures NADH dehydrogenase protein. Equal protein amounts are used with a specific monoclonal capture and detection antibodies reactive to CI isolates. All standards, samples, and controls are run in duplicate in 96-well plates pre-coated with a capture antibody provided in the Abcam kit. A standard curve prepared from the HeLa lyophilized cell extract as well as additional standards prepared from a postmortem human PFC brain extract are used. The standard CI ELISA protein curve is calculated from the regression of the average OD 450 nm reading of each HeLa standard against their concentrations. The second part of the assay is the CI activity assay. Each brain sample is diluted to the same CI protein (amount determined above by ELISA) to determine the rate of CI activity. All samples and controls are run in duplicate according to the manufacturer’s protocol (AB109721, Abcam, Cambridge, UK). The rate of activity is expressed as the change in absorbance per minute using this equation: Complex I activity for each sample (milli-optical density units, mOD/min) = (1,000 × ΔOD450/30 min) where Δ represents the final minus initial OD450.

### Immunohistochemistry (IHC) for Colocalization of Mitochondria and Synaptic Marker

Fresh frozen DLPFC tissue blocks (∼1 cm^3^) were immersion-fixed in freshly made 4% PFA (in PBS) for 24 hours at 4°C. Post-fixation, tissue blocks were cryopreserved in 20% sucrose and 0.1% Na-Azide in PBS for 3 days; then placed in 30% sucrose for 3 days at 4°C. After cryopreservation, blocks are stored frozen at -80°C until cryo-sectioning. Free-floating 10 µm tissue cryostat sections are collected in 0.1% sodium azide in PBS. Antigen-retrieval was performed through incubation of free-floating tissue sections in 1 mM EDTA buffer (pH 8) at 100°C and high pressure for 5 min. After antigen retrieval, slices were incubated in PBS-Triton (0.3%) for 1 hour. Slices were blocked with 10% goat serum. Primary antibodies for excitatory postsynaptic markers (PSD95) (MA1-045, Thermofisher) and mitochondria (TOM40) (ab185543, Abcam) were added and incubated for 24 hours at 4°C. Next day, slices were washed with PBS three times followed by respective secondary antibodies incubation for 24 hours at 4°C. After washing secondary antibodies, slices were incubated with DAPI as a nuclear counterstain for 10 min. Next, slices were incubated in 70% ethanol for 5 min followed by incubation with auto- fluorescence eliminator reagent (Sigma Aldrich, Cat#2160) for 1 min. Slides were mounted in Prolong diamond antifade solution (Thermofisher, P36970). Digital z-stack images are collected using a Zeiss LSM 780 confocal microscope with oil X 63 objective (NA=1.4). Acquired image parameters: voxel size 0.07 x 0.07 x 0.32 micron^3^, 1024 x 1024 pixels with a depth of 12 bit. Colocalization analysis was performed through IMARIS software (Bitplane). The spots of TOM40 and PSD95 with diameter of greater than 1.2 µm were excluded from the analysis. The colocalization was defined as the number of TOM40 spots within the distance of 0.25 µm from PSD95 spots. 5-8 images from 2-3 sections per subject were captured. The total number of subjects evaluated for IHC was 51.

## References

1. Whitehurst T, Howes O. The Role of Mitochondria in the Pathophysiology of Schizophrenia: A Critical Review of the Evidence Focusing on Mitochondrial Complex One. Neurosci Biobehav Rev. 2021.

2. Clay HB, Sillivan S, Konradi C. Mitochondrial dysfunction and pathology in bipolar disorder and schizophrenia. Int J Dev Neurosci. 2011;29(3):311–24.

3. Kim Y, Vadodaria KC, Lenkei Z, Kato T, Gage FH, Marchetto MC, et al. Mitochondria, Metabolism, and Redox Mechanisms in Psychiatric Disorders. Antioxid Redox Signal. 2019;31(4):275–317.

4. Chouinard VA, Kim SY, Valeri L, Yuksel C, Ryan KP, Chouinard G, et al. Brain bioenergetics and redox state measured by (31)P magnetic resonance spectroscopy in unaffected siblings of patients with psychotic disorders. Schizophr Res. 2017;187:11–16.

5. Kim SY, Cohen BM, Chen X, Lukas SE, Shinn AK, Yuksel AC, et al. Redox Dysregulation in Schizophrenia Revealed by in vivo NAD+/NADH Measurement. Schizophr Bull. 2017;43(1):197–204.

6. Scaini G, Rezin GT, Carvalho AF, Streck EL, Berk M, Quevedo J. Mitochondrial dysfunction in bipolar disorder: Evidence, pathophysiology and translational implications. Neurosci Biobehav Rev. 2016;68:694–713.

7. Goncalves VF, Cappi C, Hagen CM, Sequeira A, Vawter MP, Derkach A, et al. A Comprehensive Analysis of Nuclear-Encoded Mitochondrial Genes in Schizophrenia. Biol Psychiatry. 2018;83(9):780–89.

8. Hagen CM, Goncalves VF, Hedley PL, Bybjerg-Grauholm J, Baekvad-Hansen M, Hansen CS, et al. Schizophrenia-associated mt-DNA SNPs exhibit highly variable haplogroup affiliation and nuclear ancestry: Bi-genomic dependence raises major concerns for link to disease. PLoS One. 2018;13(12):e0208828.

9. Enwright JF, Lewis DA. Similarities in Cortical Transcriptome Alterations Between Schizophrenia and Bipolar Disorder Are Related to the Presence of Psychosis. Schizophr Bull. 2021;47(5):1442–51.

10. Glausier JR, Enwright JF, 3rd, Lewis DA. Diagnosis- and Cell Type-Specific Mitochondrial Functional Pathway Signatures in Schizophrenia and Bipolar Disorder. Am J Psychiatry. 2020;177(12):1140–50.

11. Sequeira A, Martin MV, Rollins B, Moon EA, Bunney WE, Macciardi F, et al. Mitochondrial mutations and polymorphisms in psychiatric disorders. Front Genet. 2012;3:103.

12. Mamdani F, Rollins B, Morgan L, Sequeira PA, Vawter MP. The somatic common deletion in mitochondrial DNA is decreased in schizophrenia. Schizophr Res. 2014;159(2-3):370–5.

13. Rollins BL, Morgan L, Hjelm BE, Sequeira A, Schatzberg AF, Barchas JD, et al. Mitochondrial Complex I Deficiency in Schizophrenia and Bipolar Disorder and Medication Influence. Mol Neuropsychiatry. 2018;3(3):157–69.

14. Maurer I, Zierz S, Moller H. Evidence for a mitochondrial oxidative phosphorylation defect in brains from patients with schizophrenia. Schizophr Res. 2001;48(1):125–36.

15. Kathuria A, Lopez-Lengowski K, Jagtap SS, McPhie D, Perlis RH, Cohen BM, et al. Transcriptomic Landscape and Functional Characterization of Induced Pluripotent Stem Cell-Derived Cerebral Organoids in Schizophrenia. JAMA Psychiatry. 2020;77(7):745–54.

16. Mertens J, Wang QW, Kim Y, Yu DX, Pham S, Yang B, et al. Differential responses to lithium in hyperexcitable neurons from patients with bipolar disorder. Nature. 2015;527(7576):95-9.

17. Zeppillo T, Schulmann A, Macciardi F, Hjelm BE, Focking M, Sequeira PA, et al. Functional impairment of cortical AMPA receptors in schizophrenia. Schizophr Res. 2020.

18. Hjelm BE, Rollins B, Morgan L, Sequeira A, Mamdani F, Pereira F, et al. Splice-Break: exploiting an RNA-seq splice junction algorithm to discover mitochondrial DNA deletion breakpoints and analyses of psychiatric disorders. Nucleic Acids Res. 2019;47(10):e59.

19. Curran OE, Qiu Z, Smith C, Grant SGN. A single-synapse resolution survey of PSD95- positive synapses in twenty human brain regions. Eur J Neurosci. 2021;54(8):6864–81.

20. Angrand L, Boukouaci W, Lajnef M, Richard JR, Andreazza A, Wu CL, et al. Low peripheral mitochondrial DNA copy number during manic episodes of bipolar disorders is associated with disease severity and inflammation. Brain Behav Immun. 2021;98:349–56.

21. Shivakumar V, Rajasekaran A, Subbanna M, Kalmady SV, Venugopal D, Agrawal R, et al. Leukocyte mitochondrial DNA copy number in schizophrenia. Asian J Psychiatr. 2020;53:102193.

22. Frahm T, Mohamed SA, Bruse P, Gemund C, Oehmichen M, Meissner C. Lack of age-related increase of mitochondrial DNA amount in brain, skeletal muscle and human heart. Mech Ageing Dev. 2005;126(11):1192–200.

23. Mengel-From J, Thinggaard M, Dalgard C, Kyvik KO, Christensen K, Christiansen L. Mitochondrial DNA copy number in peripheral blood cells declines with age and is associated with general health among elderly. Hum Genet. 2014;133(9):1149–59.

24. Kumar P, Efstathopoulos P, Millischer V, Olsson E, Wei YB, Brustle O, et al. Mitochondrial DNA copy number is associated with psychosis severity and anti-psychotic treatment. Sci Rep. 2018;8(1):12743.

25. Fuke S, Kametani M, Kato T. Quantitative analysis of the 4977-bp common deletion of mitochondrial DNA in postmortem frontal cortex from patients with bipolar disorder and schizophrenia. Neurosci Lett. 2008;439(2):173–7.

26. Holper L, Ben-Shachar D, Mann JJ. Multivariate meta-analyses of mitochondrial complex I and IV in major depressive disorder, bipolar disorder, schizophrenia, Alzheimer disease, and Parkinson disease. Neuropsychopharmacology. 2019;44(5):837–49.

27. Fries GR, Bauer IE, Scaini G, Valvassori SS, Walss-Bass C, Soares JC, et al. Accelerated hippocampal biological aging in bipolar disorder. Bipolar Disord. 2020;22(5):498–507.

28. Li Z, Hu M, Zong X, He Y, Wang D, Dai L, et al. Association of telomere length and mitochondrial DNA copy number with risperidone treatment response in first-episode antipsychotic-naive schizophrenia. Sci Rep. 2015;5:18553.

29. Valiente-Palleja A, Torrell H, Alonso Y, Vilella E, Muntane G, Martorell L. Increased blood lactate levels during exercise and mitochondrial DNA alterations converge on mitochondrial dysfunction in schizophrenia. Schizophr Res. 2020;220:61–68.

30. Chang CC, Jou SH, Lin TT, Liu CS. Mitochondrial DNA variation and increased oxidative damage in euthymic patients with bipolar disorder. Psychiatry Clin Neurosci. 2014;68(7):551–7.

31. Tsujii N, Otsuka I, Okazaki S, Yanagi M, Numata S, Yamaki N, et al. Mitochondrial DNA Copy Number Raises the Potential of Left Frontopolar Hemodynamic Response as a Diagnostic Marker for Distinguishing Bipolar Disorder From Major Depressive Disorder. Front Psychiatry. 2019;10:312.

32. Wang D, Li Z, Liu W, Zhou J, Ma X, Tang J, et al. Differential mitochondrial DNA copy number in three mood states of bipolar disorder. BMC Psychiatry. 2018;18(1):149.

33. Zole E, Zadinane K, Pliss L, Ranka R. Linkage between mitochondrial genome alterations, telomere length and aging population. Mitochondrial DNA A DNA Mapp Seq Anal. 2018;29(3):431–38.

34. Wachsmuth M, Hubner A, Li M, Madea B, Stoneking M. Age-Related and Heteroplasmy-Related Variation in Human mtDNA Copy Number. PLoS Genet. 2016;12(3):e1005939.

35. Sabunciyan S, Kirches E, Krause G, Bogerts B, Mawrin C, Llenos IC, et al. Quantification of total mitochondrial DNA and mitochondrial common deletion in the frontal cortex of patients with schizophrenia and bipolar disorder. J Neural Transm (Vienna). 2007;114(5):665–74.

36. Ben-Shachar D, Zuk R, Gazawi H, Reshef A, Sheinkman A, Klein E. Increased mitochondrial complex I activity in platelets of schizophrenic patients. Int J Neuropsychopharmacol. 1999;2(4):245–53.

37. Dror N, Klein E, Karry R, Sheinkman A, Kirsh Z, Mazor M, et al. State-dependent alterations in mitochondrial complex I activity in platelets: a potential peripheral marker for schizophrenia. Mol Psychiatry. 2002;7(9):995–1001.

38. Hroudova J, Fisar Z, Hansikova H, Kalisova L, Kitzlerova E, Zverova M, et al. Mitochondrial Dysfunction in Blood Platelets of Patients with Manic Episode of Bipolar Disorder. CNS Neurol Disord Drug Targets. 2019;18(3):222–31.

39. Andreazza AC, Shao L, Wang JF, Young LT. Mitochondrial complex I activity and oxidative damage to mitochondrial proteins in the prefrontal cortex of patients with bipolar disorder. Arch Gen Psychiatry. 2010;67(4):360–8.

40. Roberts RC. Mitochondrial dysfunction in schizophrenia: With a focus on postmortem studies. Mitochondrion. 2021;56:91–101.

41. Sheng ZH, Cai Q. Mitochondrial transport in neurons: impact on synaptic homeostasis and neurodegeneration. Nat Rev Neurosci. 2012;13(2):77–93.

42. Berdenis van Berlekom A, Muflihah CH, Snijders G, MacGillavry HD, Middeldorp J, Hol EM, et al. Synapse Pathology in Schizophrenia: A Meta-analysis of Postsynaptic Elements in Postmortem Brain Studies. Schizophr Bull. 2020;46(2):374–86.

43. Roberts RC, McCollum LA, Schoonover KE, Mabry SJ, Roche JK, Lahti AC. Ultrastructural evidence for glutamatergic dysregulation in schizophrenia. Schizophr Res. 2020.

44. Roberts RC, Barksdale KA, Roche JK, Lahti AC. Decreased synaptic and mitochondrial density in the postmortem anterior cingulate cortex in schizophrenia. Schizophr Res. 2015;168(1-2):543–53.

45. Aganova EA, Uranova NA. Morphometric analysis of synaptic contacts in the anterior limbic cortex in the endogenous psychoses. Neurosci Behav Physiol. 1992;22(1):59–65.

46. Uranova NA, Vostrikov VM, Vikhreva OV, Zimina IS, Kolomeets NS, Orlovskaya DD. The role of oligodendrocyte pathology in schizophrenia. Int J Neuropsychopharmacol. 2007;10(4):537–45.

47. Vikhreva OV, Rakhmanova VI, Orlovskaya DD, Uranova NA. Ultrastructural alterations of oligodendrocytes in prefrontal white matter in schizophrenia: A post-mortem morphometric study. Schizophr Res. 2016;177(1-3):28–36.

